# Revisiting deficits in threat and safety appraisal in obsessive-compulsive disorder

**DOI:** 10.1101/2022.09.01.22279518

**Authors:** Luke J. Hearne, Michael Breakspear, Ben J. Harrison, Caitlin V. Hall, Hannah S. Savage, Conor Robinson, Saurabh Sonkusare, Emma Savage, Zoie Nott, Leo Marcus, Sebastien Naze, Bjorn Burgher, Andrew Zalesky, Luca Cocchi

## Abstract

Current behavioural treatment of obsessive-compulsive disorder (OCD) is informed by fear conditioning and involves iteratively re-evaluating previously threatening stimuli as safe. However, there is limited research investigating the neurobiological response to conditioning and reversal of threatening stimuli in individuals with OCD. A clinical sample of individuals with OCD (N=45) and matched healthy controls (N=45) underwent functional Magnetic Resonance Imaging (fMRI). While in the scanner, participants completed a well-validated fear reversal task and a resting-state scan. We found no evidence for group differences in task-evoked brain activation or functional connectivity in OCD. Multivariate analyses encompassing all participants in the clinical and control groups suggested that subjective appraisal of threatening and safe stimuli were associated with a larger difference in brain activity than the contribution of OCD symptoms. In particular, we observed a brain-behaviour continuum whereby heightened affective appraisal was related to increased bilateral insula activation during the task (*r* = 0.39, *p*_FWE_ = 0.001). These findings suggest that changes in conditioned threat-related processes may not be a core neurobiological feature of OCD and encourage further research on the role of subjective experience in fear conditioning.

## Introduction

Obsessive-compulsive disorder (OCD) is characterised by recurrent, unbidden thoughts (obsessions) and/or excessive ritualistic behaviours (compulsions) (American Psychiatric Association, 2013). Exposure response prevention (ERP) is an established first-line treatment for OCD and is predicated on learning theory principles of fear conditioning and extinction (Craske et al., 2018), which involves the acquisition and reversal of stimuli-threat associations. While ERP is an effective treatment, response rates remain moderate (Reid et al., 2021; Uhre et al., 2020), potentially reflecting important individual differences in the extent to which fear conditioning mechanisms are dysfunctional in OCD (Cooper & Dunsmoor, 2021). Few studies have addressed potential brain mechanisms supporting altered threat-related processing in individuals with OCD (Apergis-Schoute et al., 2017; Cano et al., 2021; Milad et al., 2013), leaving the role of aberrant threat and safety signal processing in OCD unclear.

The brain correlates of fear conditioning and reversal are well characterised in humans and rodents (Hall et al., 2022; Laing & Harrison, 2021; Schiller et al., 2008). Regions including the anterior insular cortex, dorsal anterior cingulate cortex (dACC), and ventromedial prefrontal cortex (vmPFC) play complementary roles in the processing of threat and safety signals. Specifically, the insula and dACC have been linked to conditioned threat processing, whereas the vmPFC has a more selective role in processing safety (Battaglia et al., 2022; Fullana et al., 2018; Savage et al., 2020). Significant differences between patients with OCD and controls have been observed in resting state functional connectivity between the striatum and frontal cortices, including the vmPFC (Harrison et al., 2009, 2013). The observed differences of activity within and between these brain regions align with fronto-striatal models of OCD (Rapoport, 1990).

Despite the well-characterised brain responses and clear link to current exposure therapy, studies examining the brain underpinnings of fear conditioning and reversal in clinical OCD have been few and largely inconsistent (Cano et al., 2021). For example, the vmPFC has been associated with both under- and over-activation to threat and safety signals (Apergis-Schoute et al., 2017; Milad et al., 2013). In addition, behavioural studies have only shown moderate evidence of abnormal fear conditioning in OCD (Cooper & Dunsmoor, 2021). The reasons for these inconclusive results are unclear, but one possibility is that this variability reflects heterogeneity in the clinical presentation of OCD (van den Heuvel et al., 2009).

To revisit threat and safety reversal in OCD, we conducted a task-based neuroimaging study in a richly phenotyped clinical sample of individuals with OCD and matched controls. We focused our analyses on key cortical brain regions thought to support threat and safety processing (insula, dACC, vmPFC), and subregions of the striatum (Tian et al., 2020). In addition to task-evoked responses to threatening and safe stimuli, we studied interactions between brain regions during the task-free resting state. Finally, to complement group-wise comparisons, a multivariate approach was adopted to assess correlations between symptom severity and brain processes underpinning threat and safety reversal in OCD.

## Materials and Methods

### Participants

Fifty-eight adult participants with a clinical diagnosis of OCD and 45 controls were recruited across Australia as part of a registered randomised-controlled clinical trial (ACTRN12616001687482). Of the OCD sample, one participant was excluded due to anatomical abnormalities, one was excluded due to data corruption, and eight due to excessive head motion (see brain imaging data preprocessing section). Three participants discontinued prior to completing the MRI session, leaving 45 individuals included in the final analyses. Inclusion criteria included a clinical diagnosis of OCD for greater than 12 months and no changes in pharmaceutical treatment in the past month. The diagnosis of OCD was independently confirmed by two board-certified psychiatrists (authors M.B. and B.B.).

This OCD cohort was compared to an age-, gender- and handedness-matched control sample of 45 participants with no history of neurological or psychiatric illness (Table 1). Further inclusion criteria (which applied to all participants) included age between 18 and 50 years, no history of psychotic disorders, suicide attempts, manic episodes, seizures, neurological disorders, traumatic head injuries, substance abuse disorders, alcohol or drug misuse, as well as no contraindications to MRI.

**Table 1.**
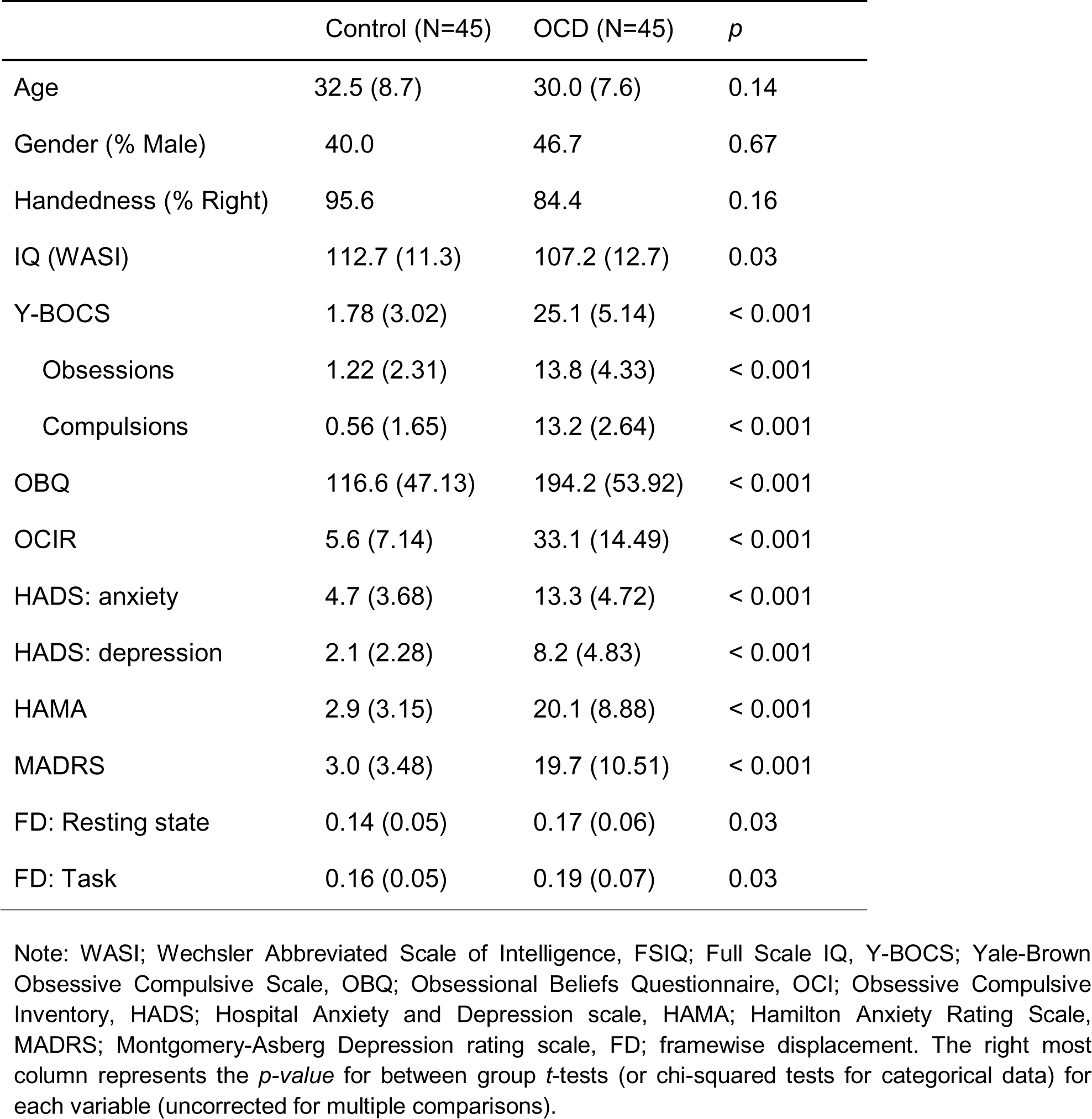
Demographics of imaging sample.

|  | Control (N=45) | OCD (N=45) | *p* |
| --- | --- | --- | --- |
| Age | 32.5 (8.7) | 30.0 (7.6) | 0.14 |
| Gender (% Male) | 40.0 | 46.7 | 0.67 |
| Handedness (% Right) | 95.6 | 84.4 | 0.16 |
| IQ (WASI) | 112.7 (11.3) | 107.2 (12.7) | 0.03 |
| Y-BOCS | 1.78 (3.02) | 25.1 (5.14) | < 0.001 |
| Obsessions | 1.22 (2.31) | 13.8 (4.33) | < 0.001 |
| Compulsions | 0.56 (1.65) | 13.2 (2.64) | < 0.001 |
| OBQ | 116.6 (47.13) | 194.2 (53.92) | < 0.001 |
| OCIR | 5.6 (7.14) | 33.1 (14.49) | < 0.001 |
| HADS: anxiety | 4.7 (3.68) | 13.3 (4.72) | < 0.001 |
| HADS: depression | 2.1 (2.28) | 8.2 (4.83) | < 0.001 |
| HAMA | 2.9 (3.15) | 20.1 (8.88) | < 0.001 |
| MADRS | 3.0 (3.48) | 19.7 (10.51) | < 0.001 |
| FD: Resting state | 0.14 (0.05) | 0.17 (0.06) | 0.03 |
| FD: Task | 0.16 (0.05) | 0.19 (0.07) | 0.03 |

Note: WASI; Wechsler Abbreviated Scale of Intelligence, FSIQ; Full Scale IQ, Y-BOCS; Yale-Brown Obsessive Compulsive Scale, OBQ; Obsessional Beliefs Questionnaire, OCI; Obsessive Compulsive Inventory, HADS; Hospital Anxiety and Depression scale, HAMA; Hamilton Anxiety Rating Scale, MADRS; Montgomery-Asberg Depression rating scale, FD; framewise displacement. The right most column represents the *p-value* for between group *t*-tests (or chi-squared tests for categorical data) for each variable (uncorrected for multiple comparisons).

Participants completed a behavioural test battery conducted by a provisional psychologist and a brain imaging session (time between sessions: mean = 4.37 days, std = 3.80). The test battery included standardised measures of OCD symptoms (type and severity), anxiety, and depression, as well as intelligence (Table 1). Overall, OCD individuals recruited in this study presented with a moderate to severe pathology (mean Y-BOCS = 25.1). The full assessment battery is detailed in Supplementary information.

The study was approved by the Human Research Ethics Committee of QIMR Berghofer. Written informed consent was obtained from all participants. All procedures contributing to this work comply with the ethical standards of the relevant national and institutional committees on human experimentation and with the Helsinki Declaration of 1975, as revised in 2008.

### Threat-safety reversal task

We used an established threat-safety reversal task (Harrison et al., 2017; Savage et al., 2020) in which a neutral stimulus (conditioned stimulus; CS) is differentially conditioned with an aversive auditory event (unconditioned stimulus; US). The CS were blue and yellow spheres presented on a black background for 2 seconds. The US consisted of a 50ms of white noise presented at 75-100dB (titrated per individual) that coterminated with the paired CS. The task had three phases: habituation, conditioning, and reversal (Fig. 1A). All three phases were acquired in a single 17-minute brain imaging session. During habituation, each sphere was presented five times without US. During conditioning, the US was presented with one of the coloured spheres but not the other, generating a CS+ and CS-. The colour of initial CS+ learned in the conditioning phase was counterbalanced across individuals. During reversal, the US - CS pairing was switched (without informing individuals). In both the conditioning and reversal phases, individuals were presented with five trials of CS+ paired with the US, ten trials of the CS+ alone, and 10 trials of the CS-. The presentation of the stimuli were pseudo randomised such that no two trial types were consecutively presented. A white fixation cross, which lasted for 12 seconds, was presented between stimulus trials.

**Figure 1.**
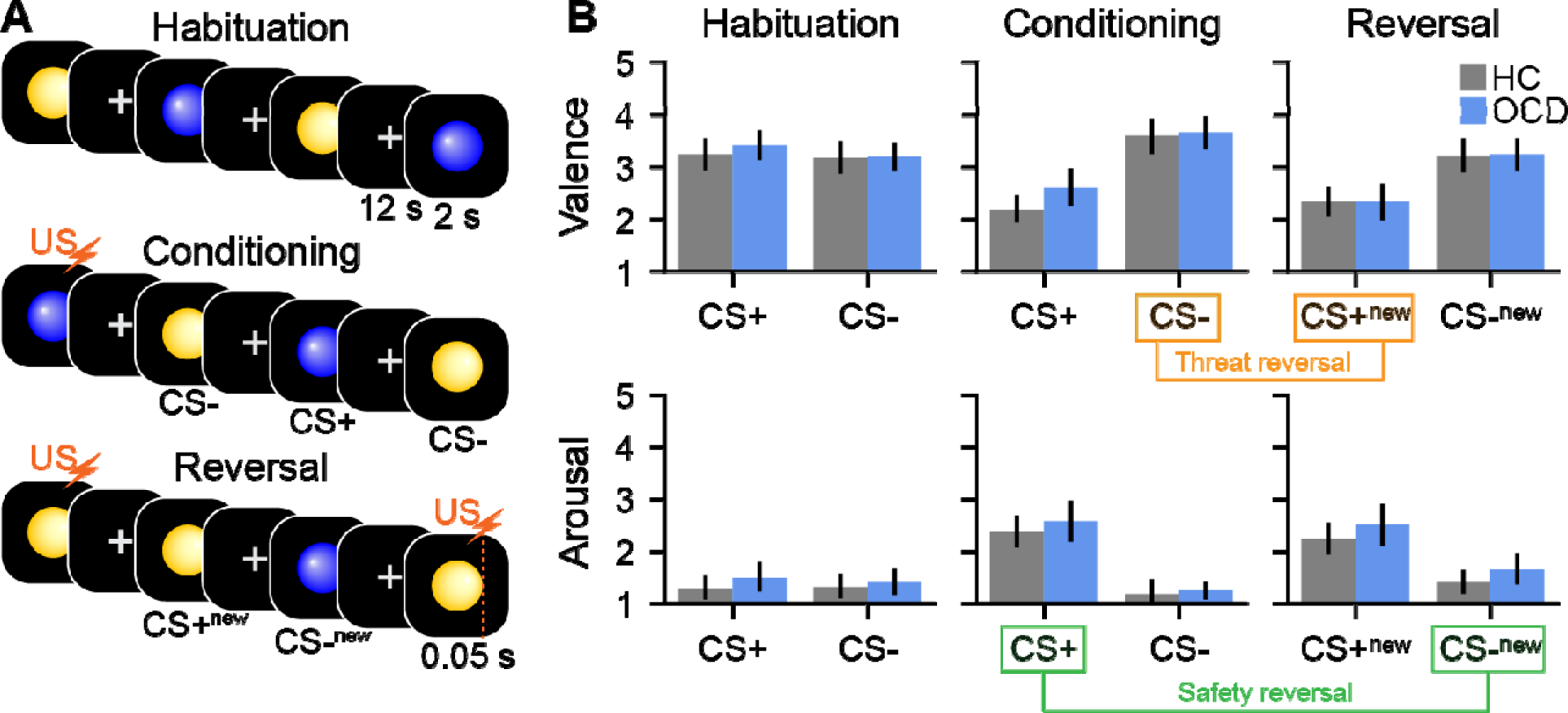
The threat-safety reversal task paradigm and subjective ratings. **A.** The experiment comprised three phases: habituation (top row), conditioning (middle row) and reversal (bottom row). Blue and yellow spheres were used as the conditioned stimuli (CS). The unconditioned stimulus (US, orange lightning bolt) was 50ms of white noise. During the conditioning phase the US co-terminated with one of the CS, forming a CS+ (threatening stimuli). Nothing was paired with the other CS, forming a CS- (safe stimuli). During the reversal phase, unknown to the participant, the US-CS pairing was switched (forming CS+_ne_ and CS-_new_. **B.** Subjective ratings corresponding to each task phase for valence (top) and arousal (bottom). Safety reversal (CS-_new_ - CS+) and threat reversal (CS+_new_ - CS-) contrasts are highlighted in green and orange, respectively. While the expected main effects of safety and threat reversal were observed, there were no differences between the healthy control (HC, grey) and OCD cohorts (blue).

Directly following completion of each phase of the task (habituation, condition, reversal), individuals were asked to rate the coloured spheres regarding anxious arousal and valence on a five-point scale (Bradley & Lang, 1994). These responses were made using a button box in the individual’s dominant hand. Individuals were familiarised with the scales and the auditory volume of the US adjusted to induce tolerable discomfort prior to the scan.

We were specifically interested in behaviour and brain activity associated with flexibly reassessing threatening stimuli as safe, and vice versa. Therefore, we contrasted the final *reversal* phase of the experiment with the second *conditioning* phase to calculate *safety reversal* (CS-^new^ > CS+) and *threat reversal* (CS+^new^ > CS-). These contrasts are identical to previous research using the same task paradigm (Savage et al., 2020) and were used in the subsequent brain activation and connectivity analyses.

For each of the arousal and valence scales, the safety and threat reversal scores (calculated as in the above paragraph) were entered in a one sample *t*-test. A between-subject *t*-test was employed to compare OCD and HC cohorts (two-tailed tests, controlling for two multiple comparisons per scale). Bayes factors for all contrasts were reported where appropriate (a default Cauchy scale factor of 0.707 was used).

At the conclusion of the task participants were asked whether they were aware of what stimuli was paired with the aversive noise in the conditioning or reversal phases of the experiment (possible answers; “a) In association with the yellow sphere, b) In association with the blue sphere, c) randomly, d) I don’t know”). Any participant who answered incorrectly at either phase of the experiment were considered unaware. Participants were also asked to rate how unpleasant they found the aversive noise on a scale of 1 to 10 (1 being “not at all unpleasant/annoying” and 10 being “extremely unpleasant/annoying”).

### Resting state

Prior to the threat-safety reversal task, a resting state acquisition was obtained. Participants viewed a black screen with a white fixation cross. They were verbally reminded to keep their eyes open, stay as still as possible, and to avoid ruminating on any specific thought.

### Brain imaging data acquisition and preprocessing

Brain imaging data were acquired on a 3T Siemens Prisma MR scanner equipped with a 64-channel head coil at the Herston Imaging Research Facility, Brisbane, Australia. Whole brain echo-planar images were acquired with the following parameters: voxel size = 2mm^3^, TR = 810 ms, multiband acceleration factor = 8, TE = 30 ms, flip angle = 53°, field of view = 212 mm, 72 slices. The fear reversal task lasted 17 minutes (1227 volumes) and the resting state was 11.9 minutes (880 volumes). Structural brain images (MPRAGE GRAPPA2) used in the preprocessing pipeline were acquired with the following parameters: voxel size = 1mm^3^, TR = 1900 ms, TE = 2.98 ms, 256 slices, flip angle = 9°. Anterior-to-posterior and posterior-to-anterior spin echo fieldmaps were acquired.

The functional brain images were preprocessed using a combination of fMRIprep (version 20.2.1) (Esteban et al., 2018), FMRIB’s ICA-based X-noiseifier (ICA-FIX) (Griffanti et al., 2014), and in-house python scripts (Supplementary information). Briefly, the data were skull stripped, corrected for susceptibility distortions, coregistered to the anatomical image and slice time corrected. Following this, denoising was conducted using ICA-FIX, which involves identifying and removing nuisance signal components using a supervised classification algorithm (Supplementary information. The data were then resampled to a standard template space (MNI152NLin2009cAsym). Data from the task paradigm were detrended and spatially smoothed with a 6 mm FWHW Gaussian filter. Resting state data were detrended, temporally filtered (0.01 - 0.1 Hz), scrubbed using a threshold of 0.5 framewise displacement (Power et al., 2012), and spatially smoothed with a 6 mm FWHW Gaussian filter. Participants with less than eight minutes of resting state data remaining after the motion scrubbing procedure were excluded (N_OCD_ = 8, see Supplementary Fig. 1).

### Task activation estimation

We used SPM12 (Wellcome Trust Centre for Neuroimaging, UK) via Nipype (Gorgolewski et al., 2011) to estimate task evoked brain activity in the threat-safety reversal contrasts. At the first level, two-second duration boxcar regressors starting at event onset (the coloured sphere stimuli) were convolved with a canonical hemodynamic response function and its temporal derivative. The model had six task regressors, representing each CS trial type across the three experiment phases: habituation, conditioning, and reversal (Fig. 1A). An additional two regressors were included to capture paired US-CS events in the conditioning and reversal phases. These regressors were modelled to control for US-related brain activity (e.g., auditory cortex activation). A final regressor representing any time point with framewise displacement above 0.5 was also included in the first level design matrix. The model was estimated using default SPM12 parameters, except for pre-whitening, which was performed using the ‘FAST’ method, due to the sub second sampling rate in the study (Bollmann et al., 2018). At the second level, two contrasts were calculated identical to the behavioural analysis: *threat* and *safety reversal* (see Threat-safety reversal task section).

#### Regions of interest

Brain regions associated with threat and safety reversal are well-characterised (Fullana et al., 2018). Thus, to maximise sensitivity, we selected brain regions *a priori*. To isolate functional regions of interest (ROI), we used statistical maps representing safety and threat reversal previously reported in Savage and colleagues (2020). These results were obtained from an independent sample of 94 individuals who completed an identical threat-safety reversal task. The maps were thresholded (*z* > 3.1) and binarised to create brain masks representing the vmPFC, PCC, bilateral insula and dACC. In addition, we added three bilateral striatal regions representing the putamen, caudate and globus pallidus based on a recent subcortical parcellation (Tian et al., 2020). In total, 11 ROI were investigated (Supplementary Fig. 2 and Supplementary Table 1).

Average regression parameter estimates (representing task-evoked brain activations) were extracted from each ROI. Two statistical tests were performed for each ROI: a two-tailed one sample *t*-test (to test replication of previous results), and a between-group two-tailed *t*-test comparing groups. Statistical significance was ascribed using *p* < 0.05 with multiple comparison correction was conducted using false discovery rate (FDR, 11 comparisons) (Benjamini & Yekutieli, 2001). As in the behavioural analysis, Bayes factors were also reported (a default Cauchy scale factor of 0.707 was used).

We also performed exploratory one-tailed, one- and two-sample whole brain analysis in SPM12 for both safety and threat reversal contrasts (one-tailed test, cluster-forming height threshold of p < 0.001, cluster thresholds of p_FDR_ < 0.01 and p_FDR_ <0.05 were tested). For group contrasts, both HC > OCD and OCD > HC contrasts were performed.

### Task and resting state connectivity estimation

OCD has also been associated with dysconnectivity between key threat and safety network brain regions (Paul et al., 2019). For the threat-safety reversal task we used the generalised psychophysiological interaction (gPPI) framework (McLaren et al., 2012) implemented via AFNI (Cox, 1996) and Nilearn (Abraham et al., 2014) to estimate FC (Supplementary information). After calculating FC matrices for each condition, the same preceding threat reversal and safety reversal contrasts were conducted (see *Threat-safety reversal task* section).

Differences in the activity of functional brain networks between healthy control and OCD have been consistently observed during the resting state (Harrison et al., 2009; Naze et al., 2023). Thus, to assess the possible context-dependence of OCD dysfunction in threat and safety networks we also investigated FC during the resting state. For the resting state data, multiple regression was used to estimate FC. For each of the 11 brain regions, all other timeseries were used as predictors in a regression model. The resulting beta coefficients were used as FC values. Multiple regression was chosen to keep the resulting FC values in the same scale as the task FC (gPPI), maximising the interpretability of the matrices. Given that the directionality of connectivity defined using regression has limited interpretability, FC values between any pair of brain regions were averaged across both directions for both the task and resting-state FC matrices.

We used the Network Based Statistic (Zalesky et al., 2010) to test for statistical significance to network components in each of the three conditions (10,000 permutations). A standard *t* threshold of 3 was used (*t* values of 2.5 and 3.5 were also tested for robustness). Two statistical tests were performed for the connectivity matrices: a one sample two-tailed t-test, and a between-group two-tailed *t*-test. For the resting state data, only the latter test was adopted.

### Multivariate analysis of brain-symptom relationships

We used partial least squares (PLS) (McIntosh & Lobaugh, 2004) to identify putative relationships between behavioural, symptom and brain responses during the task. In brief, PLS yields latent variables that maximally covary between two sets of data, here brain activations and symptoms. Non-parametric permutation and resampling methods are then used to test if these latent variables – or modes – are robust and statistically significant (Krishnan et al., 2011). Critically this approach aggregates both control and OCD cohorts in a single statistical model.

For the brain data in the PLS, we included individual brain activation parameter estimates for threat (nine ROI) and safety reversal (two ROI) contrasts. For behavioural data, we included five common symptom factors derived from the Y-BOCS: i) taboo, ii) contamination/cleaning, iii) doubts, iv) rituals/superstition, and v) hoarding/symmetry (Katerberg et al., 2010). In a second, exploratory model, we also included subjective behavioural ratings of anxious arousal and valence during the task. Principal component analysis was used to reduce the dimensionality of both the behavioural and brain data prior to the PLS analysis. This was done to ensure a more suitable number of features (five per side) for the number of observations (N=90). As the number of features was arbitrary, we conducted a control analysis to ensure this had no bearing on the results. Consistent with prior work, statistical significance of each mode was assessed with permutations (N = 20,000) and the reliability of each individual measure was assessed with bootstrapping (N = 10,000).

### Data and code availability

Analysis code is available on github (https://github.com/clinical-brain-networks/OCDbaseline_public). The whole brain statistical maps are available on neurovault (available after publication DOI).

## Results

### Group contrasts for subjective ratings of threatening and safe stimuli

We observed strong evidence of threat and safety reversal learning for both anxious arousal ratings (threat reversal; *p_FDR_* < 0.0001, safety reversal; *p_FDR_* < 0.0001) and valence ratings (threat reversal; *p_FDR_*< 0.0001, safety reversal; *p_FDR_*< 0.0001; Fig. 1B). However, there were no significant group differences for either rating scale (anxious arousal; threat reversal; *p_FDR_* > 0.99, safety reversal; *p_FDR_* > 0.99; valence; threat reversal; *p_FDR_*> 0.99, safety reversal; *p_FDR_* = 0.36, Fig. 1B, supplementary Table 2).

Bayesian statistics confirmed strong evidence of no group differences in both arousal (threat; BF_10_ = 0.30, safety; BF_10_ = 0.23) and valence (threat; BF_10_ = 0.23, safety; BF_10_ = 0.61).

Upon task completion, participants were asked whether they were aware of the pairing between specific stimuli and the aversive noise. While the entire healthy control cohort could correctly identify the conditioned stimulus in each experiment phase, 15 OCD participants could not (*X^2^*= 90.0, *p* < 0.001). Given this finding, we reanalysed the behavioural data with new grouping variables (healthy control, aware OCD, unaware OCD) and conducted ANOVAs for each of safety and threat in both the valence and arousal scales. Awareness significantly impacted subjective ratings such that the unaware group tended to demonstrate less change in ratings across experiment phases (Supplementary table 3 and Supplementary Fig. 3). In contrast, the aware OCD subgroup showed significantly higher anxious arousal in the threat reversal contrast compared to healthy controls (*t* = 2.33, *d* = 0.54, *p_FDR_* = 0.023). There was no group difference in degree of noise averseness ratings (*F* = 1.26, *n ^2^* = 0.03, *p* = 0.29).

### Group contrasts of brain activation during threat and safety reversal

Across both groups, the main effects of threat and safety reversal replicated previous task-evoked brain activation effects (Fig. 2, supplementary Table S4) (Savage et al., 2020). During safety reversal, significant effects occurred in the vmPFC and PCC (Fig. 2A): Mirroring the results of prior work, safety reversal response was characterised by reduced activation in the conditioning CS+ trials, followed by a return to baseline in the reversal CS-condition (Fig. 2B). During threat reversal, increased activation was observed in the bilateral insula, bilateral putamen and the right caudate. This threat response was characterised by an increase in activation during the CS+ in the reversal phase, compared to the CS-in the conditioning phase (Fig. 2C).

**Figure 2.**
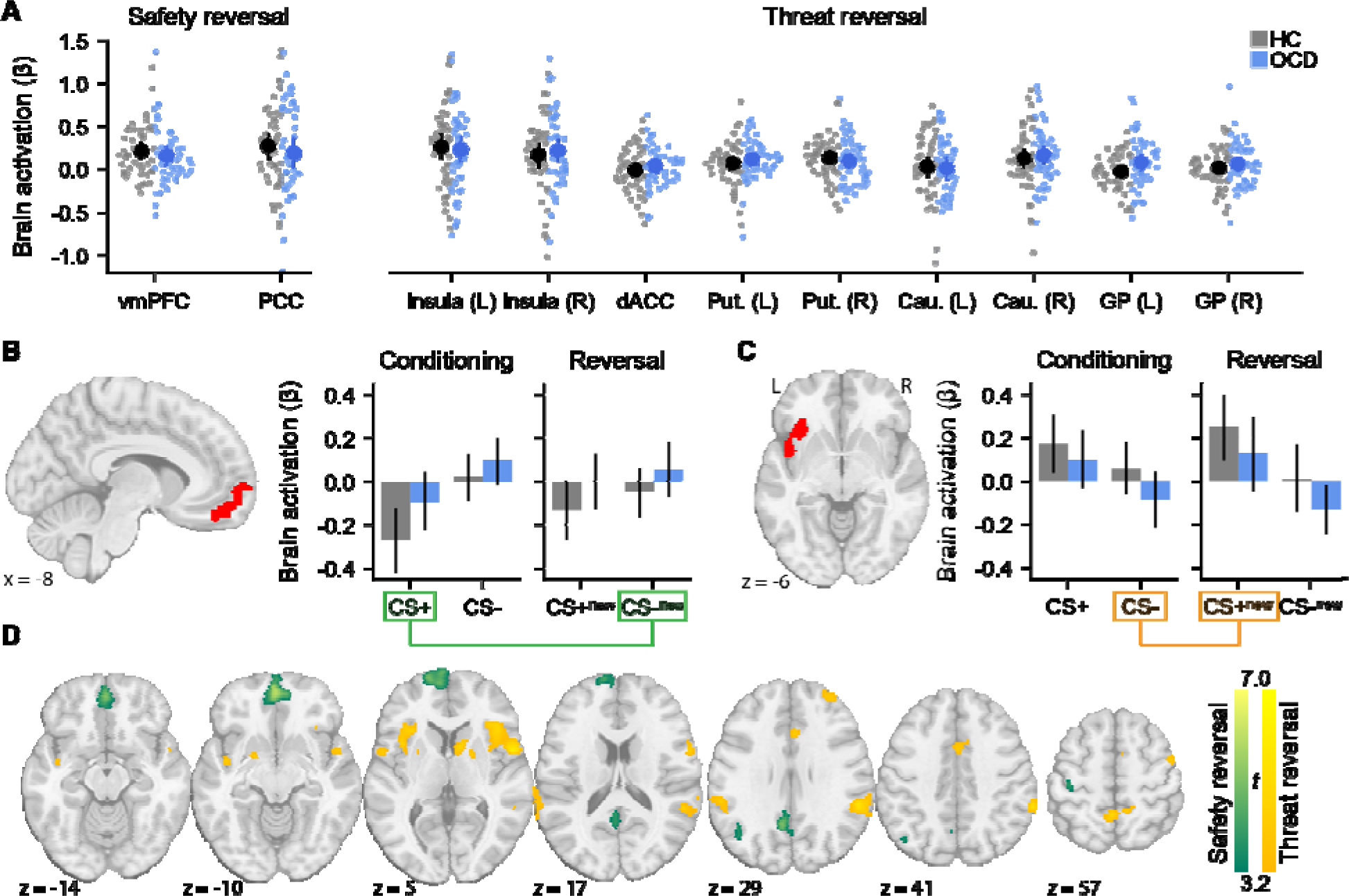
Brain activity during fear and safety reversal. Regions of interest were derived from an independent dataset (Supplementary Fig. 3). **A.** Safety (left) and threat (right) reversal activation pattern across all ROI (Supplementary Fig. 2). Raw data, as well as means and confidence intervals (95%) are displayed. There are no significant differences between healthy control (HC, grey data) and OCD (blue data) groups. **B.** VmPFC brain activation across the conditioning and reversal phases. **C.** Left insula brain activation across the conditioning and reversal phases. Safety reversal (CS-_new_ - CS+) and threat reversal (CS+_new_ - CS-) contrasts are highlighted in green and orange, respectively. **D.** Exploratory whole brain mapping results for safety (green) and threat reversal contrasts (orange) (cluster-forming height threshold: p < 0.001, cluster threshold: p_FDR_ < 0.01). No significant group differences were observed, even after relaxing the cluster threshold (p_FDR_ < 0.05). dACC; dorsal anterior cingulate cortex, vmPFC; ventromedial prefrontal cortex, PCC; posterior cingulate cortex, GP; globus pallidus, Put; putamen, Cau; caudate.

Consistent with the behavioural results, there were no significant group differences in task-evoked activation (supplementary Table 4, BF_10_ < 1 across comparisons).

A possible explanation of the null result in task-evoked patterns of brain activity is that the selected ROI were not sensitive to group differences in our clinical sample. To test, we conducted a whole-brain activation analyses (Fig. 2D, cluster-forming height threshold; p < 0.001, cluster threshold; p_FDR_ < 0.01). While the main effects of task replicated, no significant group differences were observed. However, this analysis did reveal a significant threat reversal dACC cluster in a more dorsal location than the ROI selected apriori (Savage et al., 2020).

Given the significant group differences in contingency awareness, we also tested whether awareness status was associated with differences in threat and safety brain activations. We found no evidence to support this across any of the ROI (*p_FDR_* > 0.99, see supplementary Table 5 and Fig. 4).

Most participants in the clinical sample were on pharmacological management at the time of the experiment (N_medicated_ = 35, N_unmedicated_ = 8). However, there were no statistical differences in either the behavioural or brain data when contrasting those receiving pharmacological management to those who were not.

### Group contrasts of task and resting state functional connectivity

Mirroring the task activation results, significant main effects in functional connectivity (FC) occurred for both threat (*p*_FWE_ < 0.001, Fig. 3A right) and safety reversal (*p*_FWE_ = 0.004, Fig. 3A left). Brain networks associated with these effects encompassed functional connectivity between the insula, putamen, and caudate. Safety reversal was characterised by *increased* task FC (from negative values to near zero values, Fig. 3C), whereas threat reversal was characterised by *decreased* task-related FC values (from positive to near zero, Fig. 3D). In line with the prior activation analyses, we did not detect significant group differences in task FC.

**Figure 3.**
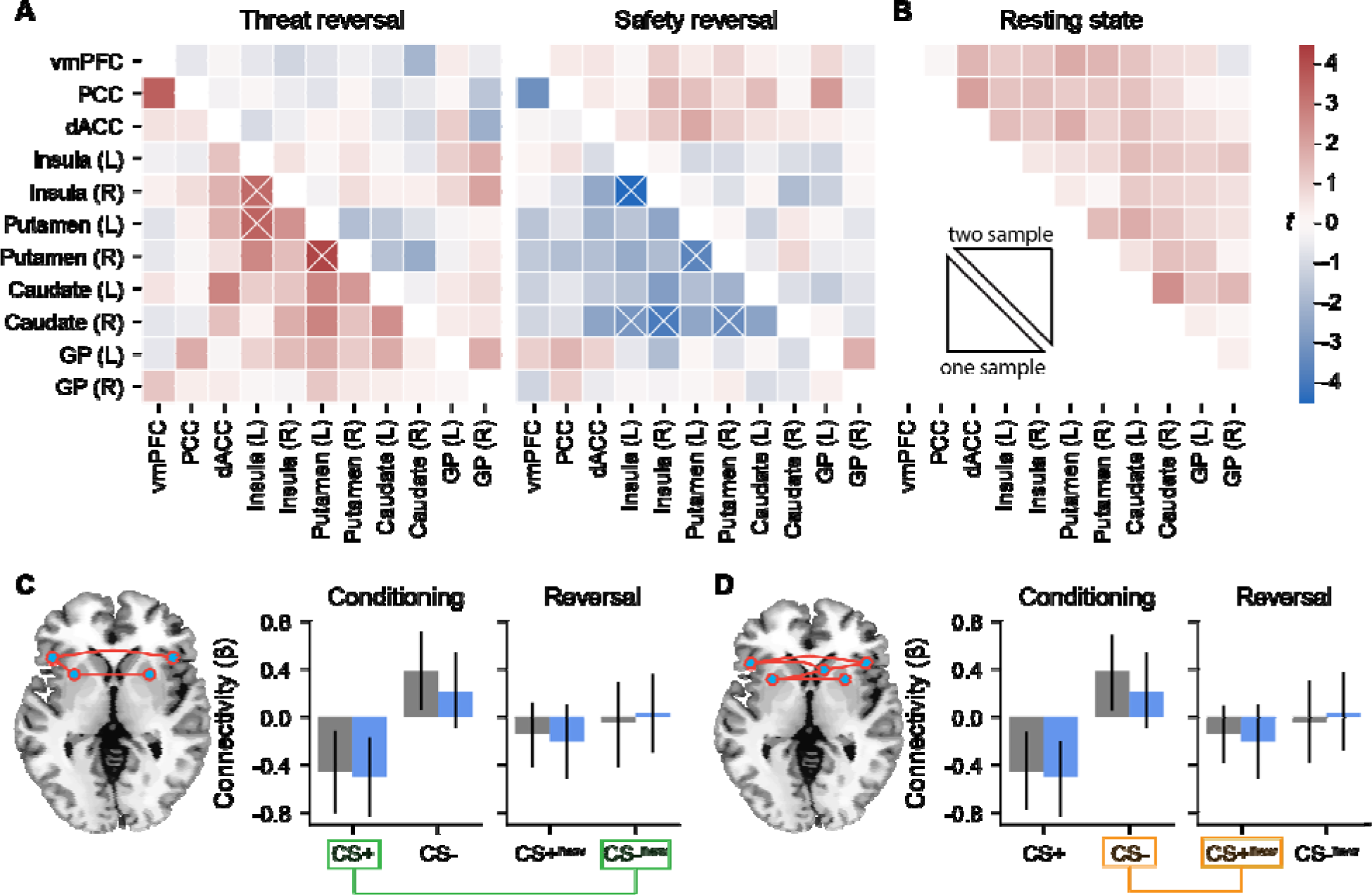
Task and resting state connectivity. **A.** Change in FC associated with threat and safety reversal. The lower triangle represents the average one-sample (across groups) response, whereas the upper triangle represents the comparison between OCD and HC. Crosses represent significant networ components (*p_FWE_* < 0.05). **B.** Differences in resting state FC. No group differences were observed. **C.** The significant safety reversal FC network component identified with the network-based statistic. Average FC values are plotted across the conditioning and reversal phases. **D.** The significant threat reversal FC network across the conditioning and reversal phases. Safety reversal (CS-_new_ - CS+) and threat reversal (CS+_new_ - CS-) contrasts are highlighted in green and orange, respectively. dACC; dorsal anterior cingulate cortex, vmPFC; ventromedial prefrontal cortex, PCC; posterior cingulate cortex, GP; globus pallidus.

No group differences in resting state FC were observed (Fig. 3B).

We also tested whether awareness was associated with differences in either task- or resting state FC. No components were detected in task FC and no significant components were detected at rest (*p_FWE_* = 0.18 contrasting healthy controls and unaware OCD, *p_FWE_* = 0.20 contrasting aware and unaware OCD). In line with the behavioural and activation results there were also no statistical differences in functional connectivity between individuals receiving pharmacological treatments and those who were not (*p_FWE_* > 0.1 when comparing unmedicated patients, medicated patients, and healthy controls, across both task and resting state contrasts). These results were robust to the choice of the network-forming threshold of *t* = 3.0).

### Multivariate mapping between task activations and behaviour

Application of PLS across both groups yielded a significant relationship between brain responses and symptom factors (*r* = 0.30, *p*_FWE_ = 0.047). A symptom profile loading on high taboo and doubts relative to other symptom categories (contamination, rituals, and hoarding) was associated with increased threat reversal-related brain activation, encompassing the bilateral putamen, insula and caudate (Fig. 4A). Given that PLS highlights a dimensional relationship, the converse brain-behaviour relationship also holds: a symptom profile of relatively low taboo and doubts was associated with decreased threat activations. It must be noted that, this brain-symptom pattern is sensitive to the number of brain and behavioural components entered in the model (Supplementary Fig. 5-6). To aid in the interpretation of the PLSC results we plotted high and low scoring individual’s raw symptom factors and brain activations in Supplementary Figure 7. This analysis revealed that individuals with relatively low taboo and doubts tended to demonstrate deactivations in the threat network.

**Figure 4.**
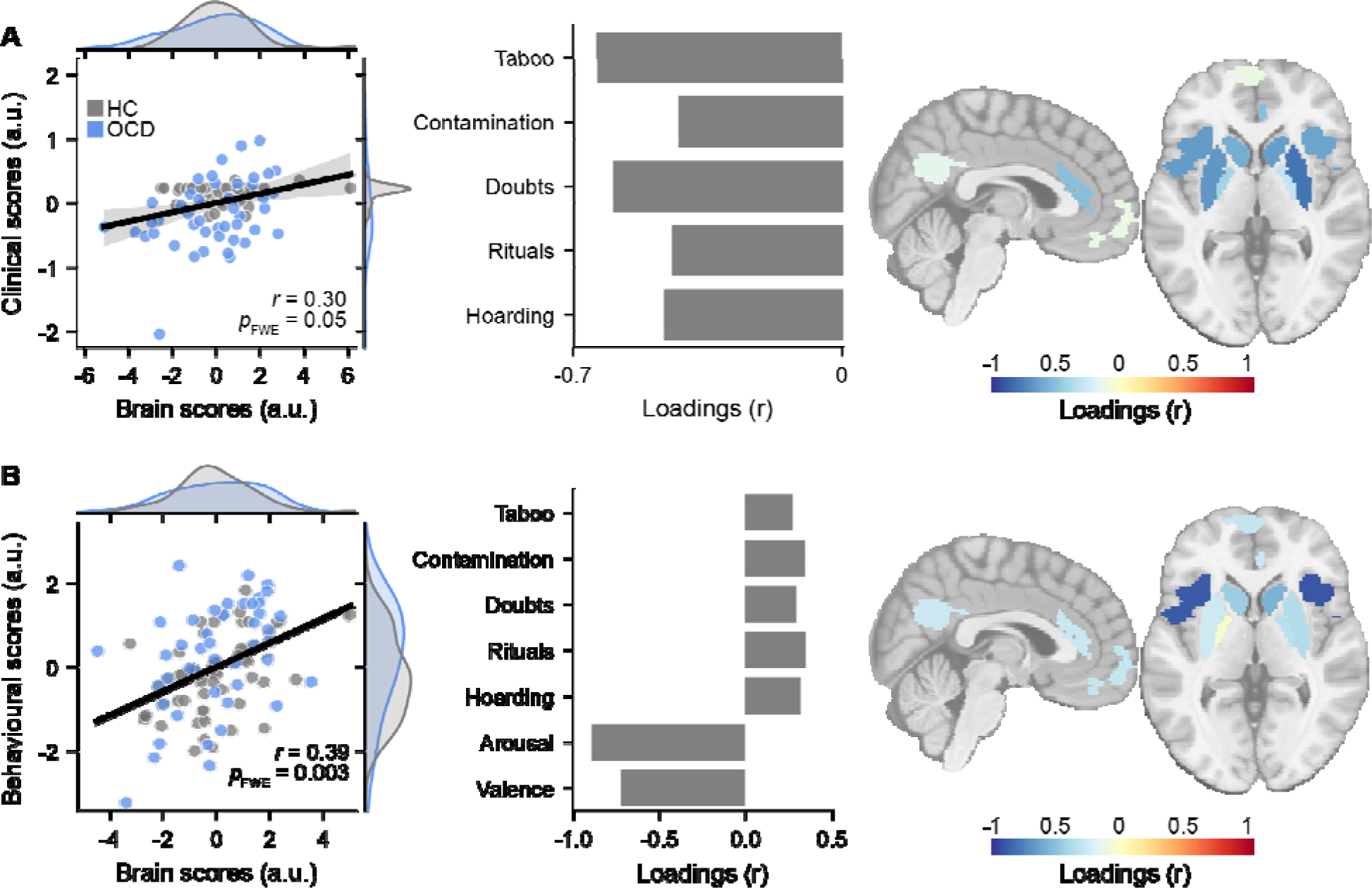
Multivariate brain-symptom relationship using partial least squares (PLS). **A.** Initial symptom factor model. The left-most panel demonstrates the relationship between individual brain activation PLSC scores and behavioural PLSC scores. Grey data represent healthy control scores, blue represent OCD scores. The middle panel shows the correlation between the original behavioural variables and the behavioural PLSC scores. The right-most panel shows the correlation between brain activation and the brain PLSC scores. The analysis highlights the association between the observed OCD symptom patterns and brain activation patterns, showing that taboo and doubt symptom factors were linked to decreased threat reversal (but not safety reversal) brain activation (and vice versa). **B.** Secondary model with arousal and valence subjective ratings included, showing that higher subjective scores were linked to increased bilateral insula activation (and vice versa).

A second exploratory PLSC model with additional subjective anxious arousal and emotional valence ratings revealed a single significant mode linking brain and behavioural data (*r* = 0.39, *p*_FWE_ = 0.001, Fig. 4B). This mode was characterised by large changes in subjective arousal and valence scores across the experiment, and to a lesser extent low Y-BOCS symptom factor scores mapping on to higher bilateral insula activation. The converse, that muted subjective ratings, and to a lesser extent, higher Y-BOCS symptom factor scores linked to lower bilateral insula activation could also be said. This effect was robust to the specific number of components chosen for the analysis (Supplementary Fig. 6). The mode was robust to removal of the participants who were unaware of the conditioned stimulus manipulation (*r* = 0.376).

## Discussion

We investigated the core brain processes supporting threat and safety responses in persons with OCD. Although we replicated the main effects of previous studies (Savage et al., 2020), there were no group differences in behaviour, brain activity or brain connectivity. Moving away from a categorical diagnostic comparison, multivariate analysis that encompassed all participants into a single cohort revealed that threat-related brain activity largely mapped onto subjective affect ratings, rather than OCD symptoms (Savage et al., 2021). These findings highlight the importance of considering subjective experience (e.g., anxious arousal and emotional valence ratings) in the assessment of fear conditioning. Our work addresses previous contradictory findings of reversal learning in OCD (Apergis-Schoute et al., 2017; Cano et al., 2021; Milad et al., 2013) by showing that the core macroscale neural circuitry underpinning both threat and safety reversal is largely unaffected when considering OCD as a relatively homogeneous clinical entity. Further analysis of resting-state data also demonstrates that a diagnosis of OCD does not map onto significant changes in the baseline activity of the threat and safety reversal systems. The clinical characteristics of our sample make it unlikely that these findings are attributed to a mild pathology given the average Y-BOCS was greater than previous studies (Apergis-Schoute et al., 2017, mean YBOCS = 21.9; Milad et al., 2013, mean YBOCS = 22.6). Additional control analyses suggested the results were not driven by ROI selection, or psychotropic medication.

Group differences were observed in *contingency awareness,* that is, whether individuals were explicitly aware of the pairing between the US and CS+ (and its reversal). The OCD cohort was significantly lower in contingency awareness, and individuals who could not identify the US – CS+ pairing tended to have muted responses to the rating scales. In contrast, persons with OCD who did demonstrate awareness showed some evidence of higher anxious arousal ratings in response to threatening stimuli compared to the healthy controls. However, as in the prior analyses, this subgrouping resulted in no significant differences in task-related brain activity or connectivity. It could be that conditioning occurred without explicit contingency awareness (Schultz and Helmstetter, 2010), specific brain regions, such as the amygdala and insula, have been shown to elicit activations regardless of whether the conditioning was subjectively reported (Knight et al., 2009). Regardless, these observations suggest future research into how subjective appraisal of threat-related stimuli might contribute to OCD psychopathology.

The clinical presentation of OCD is known to be complex and it is argued that the phenomenology is better described as a collection of partially overlapping symptom dimensions, rather than a single unitary disorder (Cervin et al., 2021; Insel et al., 2010; Mataix-Cols et al., 2005; Snorrason et al., 2021). It is plausible that this heterogeneity obscures subgroup-specific dysfunction in the processing of threat and safety (Harrison et al., 2013; Marin et al., 2020; van den Heuvel et al., 2009), leading to our null findings. Using PLS, we found evidence linking symptom factors to brain activations during fear and safety reversal. Specifically, increased subcortical and insula threat-related activations were associated with an OCD symptom profile defined by high scores on taboo and doubt symptom factors and relative to lower scores on contamination/cleaning and ritual factors. Notwithstanding the limitations of this approach and the modest effect size, our results generate new testable hypotheses regarding the potential link between patterns of threat reversal-induced brain activity and symptom dimensions in OCD.

ERP therapy, the current frontline behavioural treatment for OCD (Hirschtritt et al., 2017) is based on the hypothesis that OCD pathology is associated with core disruptions in the ability to effectively process threats and assign safety to given stimuli and thoughts. We found little neuroimaging evidence to support this. However, an important theme did emerge in our results, namely the role of subjective awareness associated with threat and safety reversal. Specifically, in a second PLSC model, we observed a relationship between threat reversal brain activations and individual differences in anxious arousal / emotional valence sensitivity, regardless of OCD status. It has been estimated that nearly half of individuals with OCD fail to show meaningful improvement following ERP therapy (Loerinc et al., 2015). These findings point towards an important role of subjective experience during fear conditioning and treatment outcome, with increased awareness likely to facilitate the therapeutic modulation of anxious state arousal and OCD symptoms.

The impact of the aversive stimulus used in the current study (a loud white noise) compared to other methods, such as electric shocks (Apergis-Schoute et al., 2017), is unclear. Given our results highlighting subjective experience, it is plausible that a patient-specific aversive stimuli may evoke larger physiological and neural differences. Likewise, we did not collect physiological measures of conditioning, such as skin conductance responses. Such measures may have helped elucidate whether conditioning occurred without subjective awareness (Mertens and Engelhard, 2020; Schultz and Helmstetter, 2010), as well as providing a concrete link to previous, related imaging studies (Apergis-Schoute et al., 2017). This represents an important limitation of the current study that should be pursued in the future.

We analysed a relatively large, single-site sample of deeply phenotyped individuals with moderate to severe OCD. However, recent work suggests that reliable brain-behaviour effects in clinical cohorts may require hundreds of participants (Libedinsky et al., 2022), with smaller cohorts - such as ours - fruitful at yielding hypotheses for further investigation. While multivariate analyses are generally more sensitive and robust than bivariate brain-wide associations, external validation in an independent cohort would strengthen confidence in our observations. Given the resource bottlenecks (of time, personnel, recruitment sources and so on), these findings also motivate the need for larger, multi-site investigations into the neurobiological mechanisms underlying the phenotype of OCD.

## Acknowledgements

The authors are indebted to the participants for generously donating their time to be part of this study.

## Author Contributions

**L.J.H:** Formal Analysis, Data Curation, Writing – Original Draft, Writing – Review & Editing, Visualization, **M.B:** Conceptualization, Investigation, Supervision, Writing – Review & Editing, **B.J.H:** Conceptualization, Writing – Review & Editing, **C.V.H:** Investigation, Data Curation, Project administration, **H.S.S:** Methodology, **C.R:** Investigation, Data Curation, Formal Analysis, **S.S:** Investigation, Data Curation, **E.S:** Investigation, Data Curation, Project administration, **Z.N:** Investigation, Data Curation, Project administration, **L.M:** Investigation, Data Curation, Project administration, **S.N:** Formal Analysis, **B.B:** Investigation, **A.Z:** Supervision, Writing – Review & Editing, **L.C:** Conceptualization, Supervision, Writing – Review & Editing, Funding Acquisition, Investigation, Formal Analysis

## References

Abraham, A., Pedregosa, F., Eickenberg, M., Gervais, P., Mueller, A., Kossaifi, J., Gramfort, A., Thirion, B., & Varoquaux, G. (2014). Machine learning for neuroimaging with scikit-learn. Frontiers in Neuroinformatics, 14.

American Psychiatric Association. (2013). Diagnostic and statistical manual of mental disorders: DSM-5. Arlington, VA.

Apergis-Schoute, A. M., Gillan, C. M., Fineberg, N. A., Fernandez-Egea, E., Sahakian, B. J., & Robbins, T. W. (2017). Neural basis of impaired safety signaling in Obsessive Compulsive Disorder. Proceedings of the National Academy of Sciences, 114(12), 3216–3221. 10.1073/pnas.1609194114

Battaglia, S., Harrison, B. J., & Fullana, M. A. (2022). Does the human ventromedial prefrontal cortex support fear learning, fear extinction or both? A commentary on subregional contributions. Molecular Psychiatry, 27(2), Article 2. 10.1038/s41380-021-01326-4

Benjamini, Y., & Yekutieli, D. (2001). The Control of the False Discovery Rate in Multiple Testing under Dependency. The Annals of Statistics, 29(4), 1165–1188.

Bollmann, S., Puckett, A. M., Cunnington, R., & Barth, M. (2018). Serial correlations in single-subject fMRI with sub-second TR. NeuroImage, 166, 152–166. 10.1016/j.neuroimage.2017.10.043

Bradley, M. M., & Lang, P. J. (1994). Measuring emotion: The self-assessment manikin and the semantic differential. Journal of Behavior Therapy and Experimental Psychiatry, 25(1), 49–59. 10.1016/0005-7916(94)90063-9

Cano, M., Martínez-Zalacaín, I., Giménez, M., Torrents-Rodas, D., Real, E., Alonso, P., Segalàs, C., Munuera, J., Menchón, J. M., Cardoner, N., Soriano-Mas, C., & Fullana, M. A. (2021). Neural correlates of fear conditioning and fear extinction and its association with cognitive-behavioral therapy outcome in adults with obsessive-compulsive disorder. Behaviour Research and Therapy, 144, 103927. 10.1016/j.brat.2021.103927

Cervin, M., Miguel, E. C., Güler, A. S., Ferrão, Y. A., Erdoğdu, A. B., Lazaro, L., Gökçe, S., Geller, D. A., Yulaf, Y., Başgül, Ş. S., Özcan, Ö., Karabekiroğlu, K., Fontenelle, L. F., Yazgan, Y., Storch, E. A., Leckman, J. F., Rosário, M. C. do, & Mataix-Cols, D. (2021). Towards a definitive symptom structure of obsessive−compulsive disorder: A factor and network analysis of 87 distinct symptoms in 1366 individuals. Psychological Medicine, 1–13. 10.1017/S0033291720005437

Cooper, S. E., & Dunsmoor, J. E. (2021). Fear conditioning and extinction in obsessive-compulsive disorder: A systematic review. Neuroscience & Biobehavioral Reviews, 129, 75–94. 10.1016/j.neubiorev.2021.07.026

Cox, R. W. (1996). AFNI: software for analysis and visualization of functional magnetic resonance neuroimages. Computers and Biomedical Research, 29(3), 162–173.

Craske, M. G., Hermans, D., & Vervliet, B. (2018). State-of-the-art and future directions for extinction as a translational model for fear and anxiety. Philosophical Transactions of the Royal Society B: Biological Sciences, 373(1742), 20170025. 10.1098/rstb.2017.0025

Esteban, O., Markiewicz, C. J., Blair, R. W., Moodie, C. A., Isik, A. I., Erramuzpe, A., Kent, J. D., Goncalves, M., DuPre, E., Snyder, M., Oya, H., Ghosh, S. S., Wright, J., Durnez, J., Poldrack, R. A., & Gorgolewski, K. J. (2018). fMRIPrep: A robust preprocessing pipeline for functional MRI. Nature Methods, 1. 10.1038/s41592-018-0235-4

Fullana, M. A., Albajes-Eizagirre, A., Soriano-Mas, C., Vervliet, B., Cardoner, N., Benet, O., Radua, J., & Harrison, B. J. (2018). Fear extinction in the human brain: A meta-analysis of fMRI studies in healthy participants. Neuroscience & Biobehavioral Reviews, 88, 16–25. 10.1016/j.neubiorev.2018.03.002

Gorgolewski, K., Burns, C., Madison, C., Clark, D., Halchenko, Y., Waskom, M., & Ghosh, S. (2011). Nipype: A Flexible, Lightweight and Extensible Neuroimaging Data Processing Framework in Python. Frontiers in Neuroinformatics, 5. https://www.frontiersin.org/articles/10.3389/fninf.2011.00013

Griffanti, L., Salimi-Khorshidi, G., Beckmann, C. F., Auerbach, E. J., Douaud, G., Sexton, C. E., Zsoldos, E., Ebmeier, K. P., Filippini, N., Mackay, C. E., Moeller, S., Xu, J., Yacoub, E., Baselli, G., Ugurbil, K., Miller, K. L., & Smith, S. M. (2014). ICA-based artefact removal and accelerated fMRI acquisition for improved resting state network imaging. NeuroImage, 95, 232–247. 10.1016/j.neuroimage.2014.03.034

Hall, C. V., Harrison, B. J., Iyer, K. K., Savage, H. S., Zakrzewski, M., Simms, L. A., Radford-Smith, G., Moran, R. J., & Cocchi, L. (2022). Microbiota links to neural dynamics supporting threat processing. Human Brain Mapping, 43(2), 733–749. 10.1002/hbm.25682

Harrison, B. J., Fullana, M. A., Via, E., Soriano-Mas, C., Vervliet, B., Martínez-Zalacaín, I., Pujol, J., Davey, C. G., Kircher, T., Straube, B., & Cardoner, N. (2017). Human ventromedial prefrontal cortex and the positive affective processing of safety signals. NeuroImage, 152, 12–18. 10.1016/j.neuroimage.2017.02.080

Harrison, B. J., Pujol, J., Cardoner, N., Deus, J., Alonso, P., López-Solà, M., Contreras-Rodríguez, O., Real, E., Segalàs, C., Blanco-Hinojo, L., Menchon, J. M., & Soriano-Mas, C. (2013). Brain Corticostriatal Systems and the Major Clinical Symptom Dimensions of Obsessive-Compulsive Disorder. Biological Psychiatry, 73(4), 321–328. 10.1016/j.biopsych.2012.10.006

Harrison, B. J., Soriano-Mas, C., Pujol, J., Ortiz, H., López-Solà, M., Hernández-Ribas, R., Deus, J., Alonso, P., Yücel, M., Pantelis, C., Menchon, J. M., & Cardoner, N. (2009). Altered Corticostriatal Functional Connectivity in Obsessive-compulsive Disorder. Archives of General Psychiatry, 66(11), 1189–1200. 10.1001/archgenpsychiatry.2009.152

Hirschtritt, M. E., Bloch, M. H., & Mathews, C. A. (2017). Obsessive-Compulsive Disorder: Advances in Diagnosis and Treatment. JAMA, 317(13), 1358–1367. 10.1001/jama.2017.2200

Insel, T., Cuthbert, B., Garvey, M., Heinssen, R., Pine, D. S., Quinn, K., Sanislow, C., & Wang, P. (2010). Research Domain Criteria (RDoC): Toward a New Classification Framework for Research on Mental Disorders. American Journal of Psychiatry, 167(7), 748–751. 10.1176/appi.ajp.2010.09091379

Katerberg, H., Delucchi, K. L., Stewart, S. E., Lochner, C., Denys, D. A. J. P., Stack, D. E., Andresen, J. M., Grant, J. E., Kim, S. W., Williams, K. A., den Boer, J. A., van Balkom, A. J. L. M., Smit, J. H., van Oppen, P., Polman, A., Jenike, M. A., Stein, D. J., Mathews, C. A., & Cath, D. C. (2010). Symptom Dimensions in OCD: Item-Level Factor Analysis and Heritability Estimates. Behavior Genetics, 40(4), 505–517. 10.1007/s10519-010-9339-z

Knight, D. C., Waters, N. S., & Bandettini, P. A. (2009). Neural substrates of explicit and implicit fear memory. NeuroImage, 45(1), 208–214. 10.1016/j.neuroimage.2008.11.015

Krishnan, A., Williams, L. J., McIntosh, A. R., & Abdi, H. (2011). Partial Least Squares (PLS) methods for neuroimaging: A tutorial and review. NeuroImage, 56(2), 455–475. 10.1016/j.neuroimage.2010.07.034

Laing, P. A. F., & Harrison, B. J. (2021). Safety learning and the Pavlovian conditioned inhibition of fear in humans: Current state and future directions. Neuroscience & Biobehavioral Reviews. 10.1016/j.neubiorev.2021.05.014

Libedinsky, I., Helwegen, K., Dannlowski, U., Fornito, A., Repple, J., Zalesky, A., Breakspear, M., van den Heuvel, M., Alzheimer’s Disease Neuroimaging Initiative, & Alzheimer’s Disease Repository Without Borders Investigators. (2022). Reproducibility of neuroimaging studies of brain disorders with hundreds-not thousands-of participants. BioRxiv.

Loerinc, A. G., Meuret, A. E., Twohig, M. P., Rosenfield, D., Bluett, E. J., & Craske, M. G. (2015). Response rates for CBT for anxiety disorders: Need for standardized criteria. Clinical Psychology Review, 42, 72–82. 10.1016/j.cpr.2015.08.004

Marin, M.-F., Hammoud, M. Z., Klumpp, H., Simon, N. M., & Milad, M. R. (2020). Multimodal Categorical and Dimensional Approaches to Understanding Threat Conditioning and Its Extinction in Individuals With Anxiety Disorders. JAMA Psychiatry, 77(6), 618. 10.1001/jamapsychiatry.2019.4833

Mataix-Cols, D., do Rosario-Campos, M. C., & Leckman, J. F. (2005). A Multidimensional Model of Obsessive-Compulsive Disorder. American Journal of Psychiatry, 162(2), 228–238. 10.1176/appi.ajp.162.2.228

McIntosh, A. R., & Lobaugh, N. J. (2004). Partial least squares analysis of neuroimaging data: Applications and advances. NeuroImage, 23, S250–S263. 10.1016/j.neuroimage.2004.07.020

McLaren, D. G., Ries, M. L., Xu, G., & Johnson, S. C. (2012). A generalized form of context-dependent psychophysiological interactions (gPPI): A comparison to standard approaches. Neuroimage, 61(4), 1277–1286.

Mertens, G., & Engelhard, I. M. (2020). A systematic review and meta-analysis of the evidence for unaware fear conditioning. Neuroscience & Biobehavioral Reviews, 108, 254–268. 10.1016/j.neubiorev.2019.11.012

Milad, M. R., Furtak, S. C., Greenberg, J. L., Keshaviah, A., Im, J. J., Falkenstein, M. J., Jenike, M., Rauch, S. L., & Wilhelm, S. (2013). Deficits in Conditioned Fear Extinction in Obsessive-Compulsive Disorder and Neurobiological Changes in the Fear Circuit. JAMA Psychiatry, 70(6), 608–618. 10.1001/jamapsychiatry.2013.914

Naze, S., Hearne, L. J., Roberts, J. A., Sanz-Leon, P., Burgher, B., Hall, C., Sonkusare, S., Nott, Z., Marcus, L., Savage, E., Robinson, C., Tian, Y. E., Zalesky, A., Breakspear, M., & Cocchi, L. (2023). Mechanisms of imbalanced frontostriatal functional connectivity in obsessive-compulsive disorder. Brain, 146(4), 1322–1327. 10.1093/brain/awac425

Paul, S., Beucke, J. C., Kaufmann, C., Mersov, A., Heinzel, S., Kathmann, N., & Simon, D. (2019). Amygdala–prefrontal connectivity during appraisal of symptom-related stimuli in obsessive–compulsive disorder. Psychological Medicine, 49(2), 278–286. 10.1017/S003329171800079X

Power, J. D., Barnes, K. A., Snyder, A. Z., Schlaggar, B. L., & Petersen, S. E. (2012). Spurious but systematic correlations in functional connectivity MRI networks arise from subject motion. NeuroImage, 59(3), 2142–2154. 10.1016/j.neuroimage.2011.10.018

Rapoport, J. L. (1990). Obsessive compulsive disorder and basal ganglia dysfunction. Psychological Medicine, 20(3), 465–469. 10.1017/S0033291700016962

Reid, J. E., Laws, K. R., Drummond, L., Vismara, M., Grancini, B., Mpavaenda, D., & Fineberg, N. A. (2021). Cognitive behavioural therapy with exposure and response prevention in the treatment of obsessive-compulsive disorder: A systematic review and meta-analysis of randomised controlled trials. Comprehensive Psychiatry, 106, 152223. 10.1016/j.comppsych.2021.152223

Savage, H. S., Davey, C. G., Fullana, M. A., & Harrison, B. J. (2020). Clarifying the neural substrates of threat and safety reversal learning in humans. NeuroImage, 207, 116427. 10.1016/j.neuroimage.2019.116427

Savage, H. S., Davey, C. G., Wager, T. D., Garfinkel, S. N., Moffat, B. A., Glarin, R. K., & Harrison, B. J. (2021). Neural mediators of subjective and autonomic responding during threat learning and regulation. NeuroImage, 245, 118643. 10.1016/j.neuroimage.2021.118643

Schiller, D., Levy, I., Niv, Y., LeDoux, J. E., & Phelps, E. A. (2008). From fear to safety and back: Reversal of fear in the human brain. The Journal of Neuroscience: The Official Journal of the Society for Neuroscience, 28(45), 11517–11525. 10.1523/JNEUROSCI.2265-08.2008

Schultz, D. H., & Helmstetter, F. J. (2010). Classical Conditioning of Autonomic Fear Responses Is Independent of Contingency Awareness. Journal of Experimental Psychology. Animal Behavior Processes, 36(4), 495–500. 10.1037/a0020263

Snorrason, I., Beard, C., Peckham, A. D., & Björgvinsson, T. (2021). Transdiagnostic dimensions in obsessive-compulsive and related disorders: Associations with internalizing and externalizing symptoms. Psychological Medicine, 51(10), 1657–1665. 10.1017/S0033291720000380

Tian, Y., Margulies, D. S., Breakspear, M., & Zalesky, A. (2020). Topographic organization of the human subcortex unveiled with functional connectivity gradients. Nature Neuroscience, 23(11), Article 11. 10.1038/s41593-020-00711-6

Uhre, C. F., Uhre, V. F., Lønfeldt, N. N., Pretzmann, L., Vangkilde, S., Plessen, K. J., Gluud, C., Jakobsen, J. C., & Pagsberg, A. K. (2020). Systematic Review and Meta-Analysis: Cognitive-Behavioral Therapy for Obsessive-Compulsive Disorder in Children and Adolescents. Journal of the American Academy of Child & Adolescent Psychiatry, 59(1), 64–77. 10.1016/j.jaac.2019.08.480

van den Heuvel, O. A., Remijnse, P. L., Mataix-Cols, D., Vrenken, H., Groenewegen, H. J., Uylings, H. B. M., van Balkom, A. J. L. M., & Veltman, D. J. (2009). The major symptom dimensions of obsessive-compulsive disorder are mediated by partially distinct neural systems. Brain, 132(4), 853–868. 10.1093/brain/awn267

Zalesky, A., Fornito, A., & Bullmore, E. T. (2010). Network-based statistic: Identifying differences in brain networks. NeuroImage, 53(4), 1197–1207. 10.1016/j.neuroimage.2010.06.041

