## Supplemental Materials for "Revisiting deficits in threat and safety appraisal in obsessive-compulsive disorder"

### Supplementary Information

#### 1. Neuropsychological battery

All participants completed two clinical assessments in a dedicated clinical room. The first assessment consisted in an unstructured clinical interview performed by a consultant psychiatrist in the team (M.B, B.B). This interview aimed to confirm the diagnosis of OCD and assess possible exclusion criteria, including substance abuse and contraindications to undertake an MRI scan. After completing this initial session, individuals started a second session with a provisional psychologist. In this session demographics were collected, as well as history of diagnoses, medications, and treatments (psychological or pharmacological). Individuals also completed a series of validated clinical scales: YBOCS (Goodman et al., 1989), Hospital Anxiety and Depression scale (Zigmond & Snaith, 1983), Obsessive Compulsive Inventory (Foa et al., 2002), Obsessional Beliefs Questionnaire (Obsessive Compulsive Cognitions Working Group, 2005), Hamilton Anxiety Rating Scale (Hamilton, 1959), and the Montgomery-Asberg Depression rating scale (Montgomery & Åsberg, 1979). The healthy control cohort completed a series of additional assessments not used in the current study (Hall et al., 2022).

#### 2. Brain imaging data preprocessing

Neuroimaging data preprocessing was performed using fMRIPrep 20.2.1 (Esteban, Blair, et al., 2018; Esteban, Markiewicz, et al., 2018), which is based on Nipype 1.5.1 (K. Gorgolewski et al., 2011; K. J. Gorgolewski et al., 2018). The output from fMRIprep has been minimally edited to reflect the preprocessing steps utilised in the current manuscript.

The T1-weighted (T1w) image was corrected for intensity non-uniformity (INU) with N4BiasFieldCorrection (Tustison et al., 2010), distributed with ANTs 2.3.3 (Avants et al., 2008), and used as T1w-reference throughout the workflow. The T1w-reference was then skull-stripped with a Nipype implementation of the antsBrainExtraction.sh workflow (from ANTs), using OASIS30ANTs as the target template. Brain tissue segmentation of cerebrospinal fluid (CSF), white-matter (WM) and gray-matter (GM) was performed on the brain-extracted T1w using fast (FSL 5.0.9, Zhang et al., 2001). Brain surfaces were reconstructed using recon-all (Freesurfer 6.0.1, Dale et al., 1999), and the brain mask estimated previously was refined with a custom variation of the method to reconcile ANTs-derived and FreeSurfer-derived segmentations of the cortical gray-matter of Mindboggle (Klein et al., 2017). Volume-based spatial normalisation to two standard spaces (MNI152NLin2009cAsym, MNI152NLin6Asym) was performed through nonlinear registration with antsRegistration (ANTs 2.3.3), using brain-extracted versions of both T1w reference and the T1w template. The following template were selected for spatial normalization: ICBM 152 Nonlinear Asymmetrical template version 2009c (TemplateFlow ID: MNI152NLin2009cAsym) (Fonov et al., 2009).

For both the fear conditioning and resting state BOLD runs the following preprocessing was performed. First, a reference volume and its skull-stripped version were generated using a custom methodology of fMRIPrep. A B0-nonuniformity map (or fieldmap) was estimated based on two (or more) echo-planar imaging (EPI) references with opposing phase-encoding directions, with 3dQwarp Cox and Hyde (1997) (AFNI 20160207). Based on the estimated susceptibility distortion, a corrected EPI (echo-planar imaging) reference was calculated for a more accurate co-registration with the anatomical reference. The BOLD reference was then co-registered to the T1w reference using bbregister (FreeSurfer) which implements boundary-based registration (Greve & Fischl, 2009). Co-registration was configured with six degrees of freedom. Head-motion parameters with respect to the BOLD reference (transformation matrices, and six corresponding rotation and translation parameters) are estimated before any spatiotemporal filtering using mcflirt (FSL 5.0.9, Jenkinson et al., 2002). BOLD runs were slice-time corrected using 3dTshift from AFNI 20160207. The BOLD time-series were resampled onto the following surfaces (FreeSurfer reconstruction nomenclature): fsaverage. The BOLD time-series (including slice-timing correction when applied) were resampled onto their original, native space by applying a single, composite transform to correct for head-motion and susceptibility distortions. These resampled BOLD time-series will be referred to as preprocessed BOLD in original space, or just preprocessed BOLD.

Following this standard fMRIprep preprocessing, the FIX-ICA denoising procedure was performed (described in the next section). The fMRIprep pipeline was then continued with the newly denoised data. The BOLD time-series were resampled into standard space, generating a preprocessed BOLD run in MNI152NLin2009cAsym space. First, a reference volume and its skull-stripped version were generated using a custom methodology of fMRIPrep. As typical in fMRIprep several confounding time-series were calculated based on the preprocessed BOLD. Framewise displacement was used for scrubbing and exclusion of participants (described in the main text).

Many internal operations of fMRIPrep use Nilearn 0.6.2 (Abraham et al., 2014) (mostly within the functional processing workflow. For more details of the pipeline, see the section corresponding to workflows in fMRIPrep’s documentation.

#### 3. FIX-ICA denoising

Both resting-state and task data from 26 subjects were used to train the FIX classifier. The selected subjects varied in group (13 controls and 13 OCD), gender and degree of head motion. Two raters (L.J.H. and C.R.) underwent training to identify noise components in the data in the first six subject datasets (12 scans in total). A third rater (L.C.) was consulted on any discrepancies in component labelling within this subset of the training data. Inter-rater reliability in this subset of the data was high across the two raters (average across 12 scans; percentage agreement = 96.49%, κ = 0.86). L.J.H. and C.R. went on to manually classify the remaining datasets independently. Leave-one-out cross validation was used to assess the reliability of noise component identification. When using all hand labelled data (40 scans in total), high levels of prediction accuracy were achieved across several component thresholds. We chose to use a component threshold of ten, based on good performance in the current dataset (true positive rate = 96.78%, true negative rate = 91.55%) and typical thresholds used in the literature (Griffanti et al., 2014; Salimi-Khorshidi et al., 2014). The trained classifier was applied to all participants and noise components, and 24 motion parameters were removed from the data using the recommended ‘non-aggressive’ regression approach.

#### 4. Psycho-physiological interactions

Psycho-physiological interaction (PPI) analysis was performed using a combination of Nilearn (Abraham et al., 2014) and AFNI (Cox, 1996). Timeseries from each region of interest were extracted and detrended. For each ROI a separate PPI regression analysis was performed, where each ROI was considered the 'seed' and all other timeseries were the targets. Each regression model included the seed timeseries, task onsets convolved with the hemodynamic response function and the psycho-physiological interaction term. Coefficients from each regression model corresponding to the PPI term filled a single row of the resulting connectivity matrix in each condition.

AFNI was used to create the PPI regressor. Each seed timeseries was upsampled (*1dUpsample*) deconvolved (*3dTfitter*) creating the 'physiological' timeseries. For each condition, the 'psychological' timeseries was created identically to a typical brain activity regressor, by convolving task onsets with a hemodynamic response function, followed by upsampling. These two timeseries were used to create the psychophysical interaction term, which was subsequently reconvolved with the hemodynamic response function and downsampled into the original timing.

#### 5. Partial least squares correlation

Partial least squares correlation (PLSC) was used to relate brain and behavioural data in a multivariate framework (McIntosh & Lobaugh, 2004). The brain activation and behavioural data were organised in two matrices: **X** (subject x brain activity, size 90 x 11) and **Y** (subject x behaviour, size 90 x 5). **X** included threat reversal activation estimates in 9 ROI (bilateral insula, putamen, caudate, globus pallidus and the ACC), and safety reversal activation estimates for the vmPFC and PCC. In the initial model, **Y** included symptom data only. The YBOCS-derived symptom factors were calculated by weighting and summing item-level YBOCS responses by the weights and categories described in Katerberg and colleagues (Katerberg et al., 2010). In the second model, task ratings were included. For each rating scale (valence and anxious arousal) the safety and threat contrasts were calculated and subtracted from one another.

To ensure an appropriate number of features were entered into the PLSC, principle components analysis (PCA) was performed as a data reduction method. The columns in both **X** and **Y** were z-scored columnwise and submitted to the PCA. In the main text were report the result of 5 PCA components per input matrix, but we also tested across a number of component sizes to ensure the final results were robust to this analysis choice.

The brain-behaviour covariance matrix was computed (**Y’X**). The resulting matrix was then subjected to singular value decomposition (SVD) (Eckart & Young, 1936).

**Y’X** = **USV**′

The singular value decomposition produced a set of mutually orthogonal latent variables, or modes. The number of latent variables (***k***) is equal to the rank of the covariance matrix, which, in this case, is nine. **U** represents the matrix of left singular vectors containing the weights, or *saliences,* for matrix **X**, **V** represents the matrix of right singular vectors containing the weights for matrix **Y,** and **S** is the diagonal matrix of singular values. The columns of **U** and **V** weight the original brain and behavioural data matrices (**X** and **Y**) so that they maximally covary. The effect size of each latent variable pairing (e.g., **U_k=1_** and **V_k=1_**) can be estimated as the ratio of the squared singular value associated with the current latent pairing to the sum of all squared singular values in **S** (McIntosh & Lobaugh, 2004). Individual brain and behavioural scores were calculated by projecting the brain and behavioural weights (**U** and **V**) onto the individual patient data.

Statistical significance of each latent variable was assessed with 20,000 permutation tests. In each permutation observations in **X** were shuffled across participants, severing the link between behavioural and brain features. The permuted data matrices were submitted to the same PLSC procedure described above to build a null distribution of singular values that could be compared to the original, non-permuted singular value. Thus, the *p*-value represents the probability of finding the size of the ‘real’ relationship under the null hypothesis that there is no meaningful relationship between the brain and behaviour.

Bootstrap resampling (N = 10,000) was used to assess the reliability of individual variables in the analysis. Participants were randomly sampled with replacement. These new data matrices, as above with the permutation procedure, were submitted to the same PLSC analysis as the original data. The bootstrap distribution was then used to generate standard errors and confidence intervals for each individual weight. The ratio between each weight and its bootstrap-estimated standard error were calculated, highlighting weights that were both large and reliable. This ratio is approximately equivalent to a z-score. Note that the procedure is standard, having been described in prior studies (Mišić et al., 2016) and is widely available as part of a software toolbox (<https://www.rotman-baycrest.on.ca/index.php?section=84>).

In our case, PCA components were used in the PLSC, making the resulting individual variable standard errors and confidence intervals difficult to interpret. To assist with this interpretation, we correlated the original data with the final latent variables scores in the main text. For completeness, we also present the PCA component weights and reliability analyses in the supplementary figures.

### Supplementary tables

**Table S1.** Region of interest centroids

| Label | x | y | z |
| --- | --- | --- | --- |
| Insula (left) | 41 | 19 | -3 |
| Insula (right) | -39 | 16 | -2 |
| dACC | 0 | 30 | 16 |
| PCC | -1 | -57 | 24 |
| vmPFC | -3 | 56 | -10 |
| Putamen (left) | -24 | 0 | 0 |
| Putamen (right) | 26 | 0 | 0 |
| Caudate (left) | -12 | 10 | 10 |
| Caudate (right) | 14 | 10 | 10 |
| Globus pallidus (left) | -18 | -4 | -2 |
| Globus pallidus (right) | 20 | -4 | -2 |

Note: dACC; dorsal anterior cingulate cortex, vmPFC; ventromedial prefrontal cortex, PCC; posterior cingulate cortex

**Table S2.** Subjective task rating statistics

| Measure | Contrast | *t* | DOF | *d* | *p_FDR_* | BF10 |
| --- | --- | --- | --- | --- | --- | --- |
| One sample *t*-test |  |  |  |  |  |  |
| Arousal | Safety reversal | -8.92 | 89 | 0.94 | < 0.0001 | > 1000 |
|  | Threat reversal | 9.51 | 89 | 1.00 | < 0.0001 | > 1000 |
| Valence | Safety reversal | 6.39 | 89 | 0.67 | < 0.0001 | > 1000 |
|  | Threat reversal | -8.95 | 89 | 0.94 | < 0.0001 | > 1000 |
| Two sample *t*-test |  |  |  |  |  |  |
| Arousal | Safety reversal | -0.21 | 88 | 0.04 | > 0.99 | 0.23 |
|  | Threat reversal | -0.81 | 88 | 0.17 | > 0.99 | 0.30 |
| Valence | Safety reversal | 1.52 | 88 | 0.32 | 0.36 | 0.61 |
|  | Threat reversal | 0.23 | 88 | 0.05 | > 0.99 | 0.23 |

Note: DOF; degrees of freedom. Multiple comparison correction was performed using FDR (Benjamini & Yekutieli, 2001).

**Table S3.** Subjective task ratings by awareness status

|  | Measure | Contrast | *F/t* | DOF | *N_p_^2^/d* | *p* | BF10 |
| --- | --- | --- | --- | --- | --- | --- | --- |
| ANOVA | Arousal | Safety reversal | 5.34 | 87 | 0.11 | 0.007 |  |
| HC v. OCD-aware |  |  | 1.10 | 47.12 | 0.28 | 0.278 | 0.407 |
| HC v. OCD-unaware |  |  | -4.06 | 46.33 | 0.90 | < 0.001 | 152.236 |
| OCD-aware v. OCD-unaware |  |  | -3.87 | 40.51 | 0.94 | < 0.001 | 71.611 |
| ANOVA |  | Threat reversal | 6.33 | 87 | 0.13 | 0.003 |  |
| HC v. OCD-aware |  |  | -2.33 | 65.17 | 0.54 | 0.023 | 2.405 |
| HC v. OCD-unaware |  |  | 1.86 | 25.58 | 0.54 | 0.074 | 1.18 |
| OCD-aware v. OCD-unaware |  |  | 3.58 | 28.14 | 1.13 | 0.001 | 34.46 |
| ANOVA | Valence | Safety reversal | 6.33 | 87 | 0.07 | 0.047 |  |
| HC v. OCD-aware |  |  | 0.47 | 58.07 | 0.11 | 0.639 | 2.405 |
| HC v. OCD-unaware |  |  | 3.05 | 32.03 | 0.79 | 0.005 | 1.18 |
| OCD-aware v. OCD-unaware |  |  | 2.25 | 38.80 | 0.63 | 0.03 | 34.46 |
| ANOVA |  | Threat reversal | 6.33 | 87 | 0.9 | 0.020 |  |
| HC v. OCD-aware |  |  | 1.64 | 72.80 | 0.36 | 0.106 | 2.405 |
| HC v. OCD-unaware |  |  | -1.65 | 23.32 | 0.50 | 0.113 | 1.18 |
| OCD-aware v. OCD-unaware |  |  | -2.79 | 20.57 | 1.00 | 0.011 | 34.46 |

**Table S4.** Region of interest statistics

|  | ROI | Contrast | *t* | DOF | *d* | *p_FDR_* | BF10 |
| --- | --- | --- | --- | --- | --- | --- | --- |
| *One sample t-test* | | | | | | | |
|  | Insula (left) | Threat reversal | 4.86 | 89 | 0.51 | < 0.001 | 3165.23 |
|  | Insula (right) | Threat reversal | 4.17 | 89 | 0.44 | < 0.001 | 265.01 |
|  | dACC | Threat reversal | 0.62 | 89 | 0.06 | > 0.99 | 0.14 |
|  | vmPFC | Safety reversal | 5.34 | 89 | 0.56 | < 0.001 | 19730.0 |
|  | PCC | Safety reversal | 4.03 | 89 | 0.42 | < 0.001 | 162.52 |
|  | Putamen (left) | Threat reversal | 3.5 | 89 | 0.37 | 0.004 | 31.27 |
|  | Putamen (right) | Threat reversal | 4.14 | 89 | 0.44 | < 0.001 | 234.44 |
|  | Caudate (left) | Threat reversal | 0.52 | 89 | 0.05 | > 0.99 | 0.13 |
|  | Caudate (right) | Threat reversal | 3.72 | 89 | 0.39 | 0.002 | 61.45 |
|  | Glob. pal. (left) | Threat reversal | 0.96 | 89 | 0.1 | > 0.99 | 0.18 |
|  | Glob. pal. (right) | Threat reversal | 1.45 | 89 | 0.15 | 0.643 | 0.32 |
| *Two sample t-test (between groups comparison)* | | | | | | | |
|  | Insula (left) | Threat reversal | 0.29 | 88 | 0.06 | < 0.99 | 0.23 |
|  | Insula (right) | Threat reversal | -0.54 | 88 | 0.11 | < 0.99 | 0.25 |
|  | dACC | Threat reversal | -0.96 | 88 | 0.2 | < 0.99 | 0.33 |
|  | vmPFC | Safety reversal | 0.9 | 88 | 0.19 | 0.697 | 0.32 |
|  | PCC | Safety reversal | 0.73 | 88 | 0.15 | 0.697 | 0.28 |
|  | Putamen (left) | Threat reversal | -0.86 | 88 | 0.18 | < 0.99 | 0.3 |
|  | Putamen (right) | Threat reversal | 0.67 | 88 | 0.14 | < 0.99 | 0.27 |
|  | Caudate (left) | Threat reversal | 0.26 | 88 | 0.05 | < 0.99 | 0.23 |
|  | Caudate (right) | Threat reversal | -0.44 | 88 | 0.09 | < 0.99 | 0.24 |
|  | Glob. pal. (left) | Threat reversal | -1.82 | 88 | 0.38 | < 0.99 | 0.94 |
|  | Glob. pal. (right) | Threat reversal | -0.87 | 88 | 0.18 | < 0.99 | 0.31 |

Note: ROI; region of interest, DOF; degrees of freedom, dACC; dorsal anterior cingulate cortex, vmPFC; ventromedial prefrontal cortex, PCC; posterior cingulate cortex, Glob. pal.; globus pallidus.

**Table S5.** Impact of awareness on brain activation

|  | ROI | Contrast | *F* | DOF | *n_p_^2^* | *p_FDR_* |
| --- | --- | --- | --- | --- | --- | --- |
|  | Insula (left) | Threat reversal | 0.81 | 2/87 | 0.02 | > 0.99 |
|  | Insula (right) | Threat reversal | 0.27 | 2/87 | 0.01 | > 0.99 |
|  | dACC | Threat reversal | 0.65 | 2/87 | 0.01 | > 0.99 |
|  | vmPFC | Safety reversal | 0.41 | 2/87 | 0.01 | > 0.99 |
|  | PCC | Safety reversal | 0.33 | 2/87 | 0.01 | > 0.99 |
|  | Putamen (left) | Threat reversal | 0.8 | 2/87 | 0.02 | > 0.99 |
|  | Putamen (right) | Threat reversal | 0.66 | 2/87 | 0.01 | > 0.99 |
|  | Caudate (left) | Threat reversal | 0.07 | 2/87 | < 0.01 | > 0.99 |
|  | Caudate (right) | Threat reversal | 0.1 | 2/87 | < 0.01 | > 0.99 |
|  | Glob. pal. (left) | Threat reversal | 2.41 | 2/87 | 0.05 | > 0.99 |
|  | Glob. pal. (right) | Threat reversal | 0.61 | 2/87 | 0.01 | > 0.99 |

### Supplementary figures

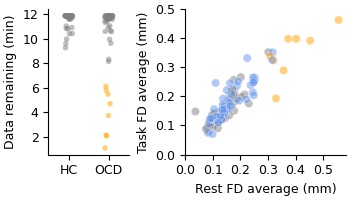

**Figure S1.** Head motion exclusions. Participants were excluded based on the amount of data remaining after head motion scrubbing in the resting state scan. Orange data represent excluded participants, blue represent the OCD cohort, and grey represent healthy controls. The left panel shows the amount of data remaining after the head motion scrubbing procedure in the resting state scan. Any participants with less than eight minutes of data remaining were excluded. The right panel shows the relationship between average head motion during the task versus during the resting state. Criteria were based on the resting state data given the impact of head motion is more severe in resting state functional connectivity analyses.

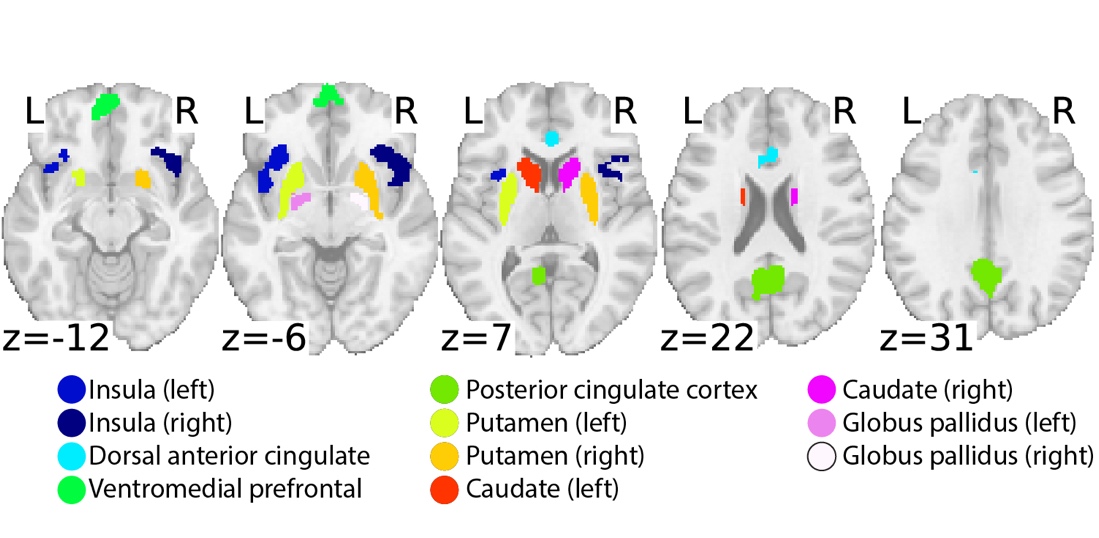

**Figure S2.** Regions of interest identified from Savage and colleagues (2020) and Tian and colleagues (2020). In total, eleven regions of interest were used in the main analyses.

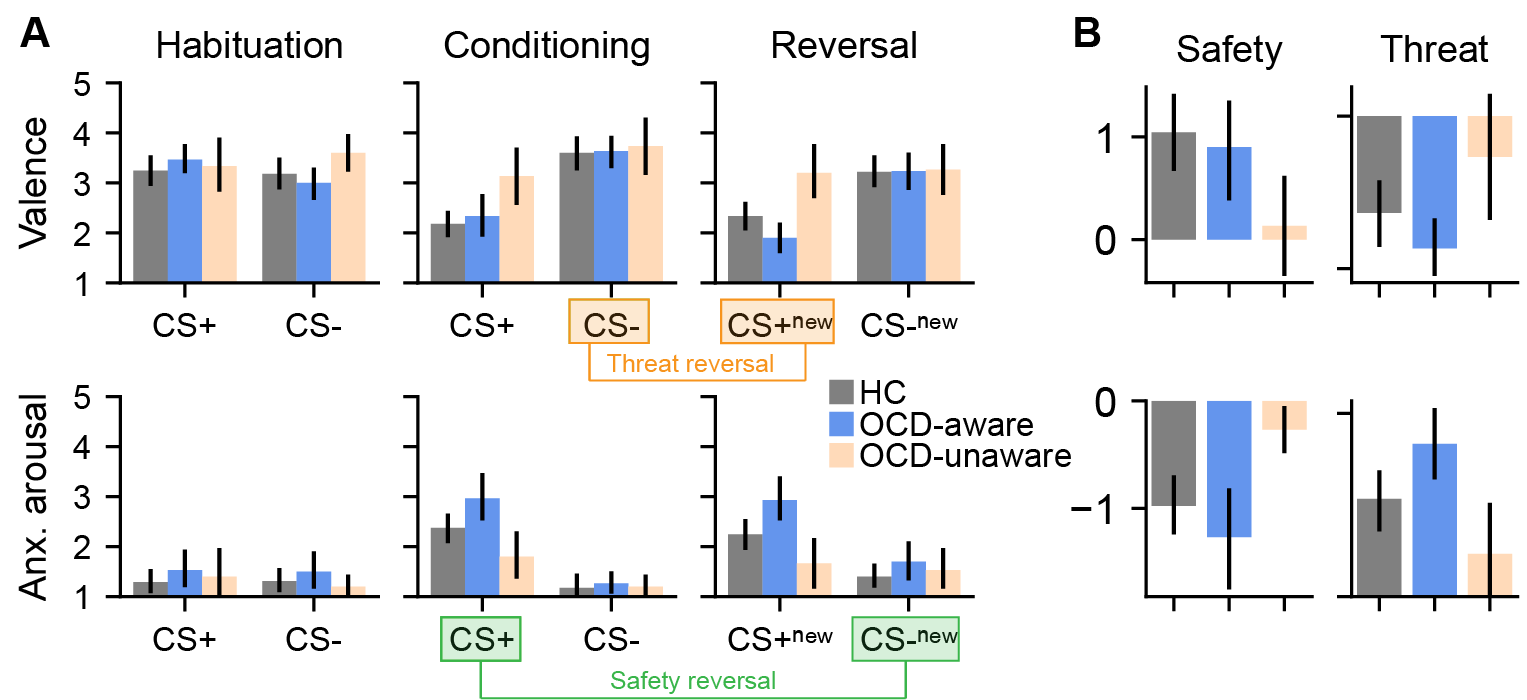

**Figure S3.** Threat-safety reversal task subjective ratings by awareness. **A.** Subjective ratings corresponding to each task phase for valence (top) and arousal (bottom). Aware (blue) and unaware (peach) OCD cohorts are plotted separately. Safety reversal (CS-_new_ - CS+) and threat reversal (CS+_new_ - CS-) contrasts are highlighted in green and orange, respectively. **B.** Subjective ratings in the safety and threat reversal contrasts. Unaware participants tended to show little change in ratings across phases.

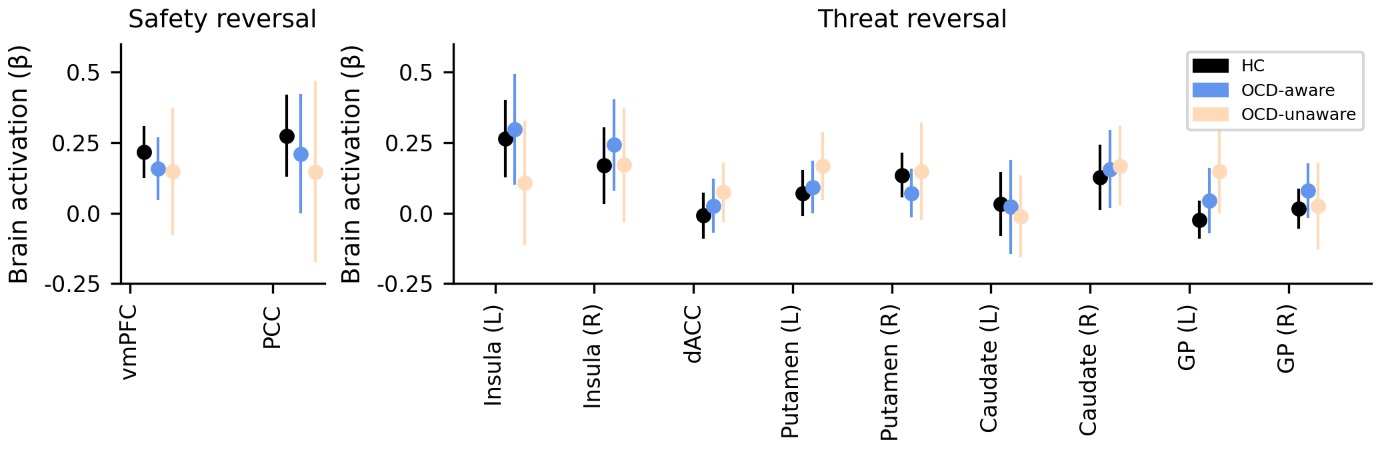

**Figure S4.** Brain activity during fear and safety reversal across awareness contingencies. Mean and 95% confidence intervals are plotted (black = HC, blue = OCD and aware, peach = OCD and unaware). No between-group statistical differences were observed across any of the regions of interest.

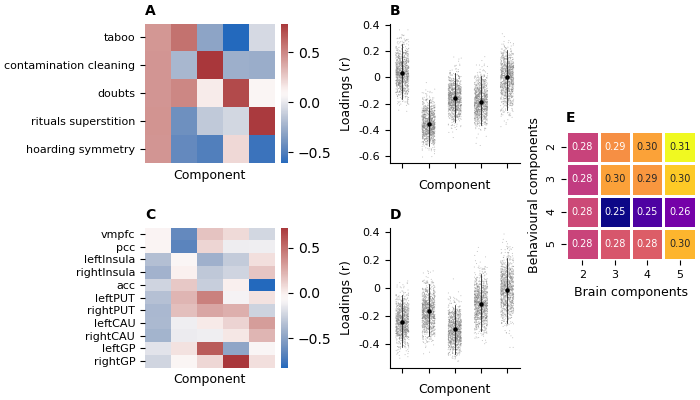

**Figure S5.** Supplementary results for the PLSC with symptoms only. **A.** PCA component weightings for the symptom factors entered in the analysis **B.** Subsequent PCA – PLSC loading patterns (black lines indicate 95% confidence intervals and grey dots indicate bootstraps). **C.** PCA component weightings for the brain activations entered in the analysis. **D.** Subsequent PCA – PLSC loading patterns (black lines indicate 95% confidence intervals and grey dots indicate bootstraps). **E.** Results of varying the number of PCA components entered in the PLSC analysis. In this case, the final correlation between brain and behaviour was somewhat unstable.

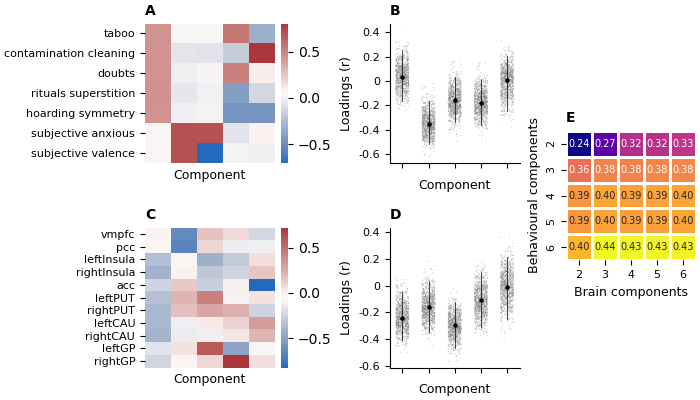

**Figure S6**. Supplementary results for the secondary PLSC with additional subjective ratings. **A.** PCA component weightings for the symptom factors entered in the analysis **B.** Subsequent PCA – PLSC loading patterns (black lines indicate 95% confidence intervals and grey dots indicate bootstraps). **C.** PCA component weightings for the brain activations entered in the analysis. **D.** Subsequent PCA – PLSC loading patterns (black lines indicate 95% confidence intervals and grey dots indicate bootstraps). **E.** Results of varying the number of PCA components entered in the PLSC analysis. The final correlation between brain and behaviour was more stable than the previous analysis.

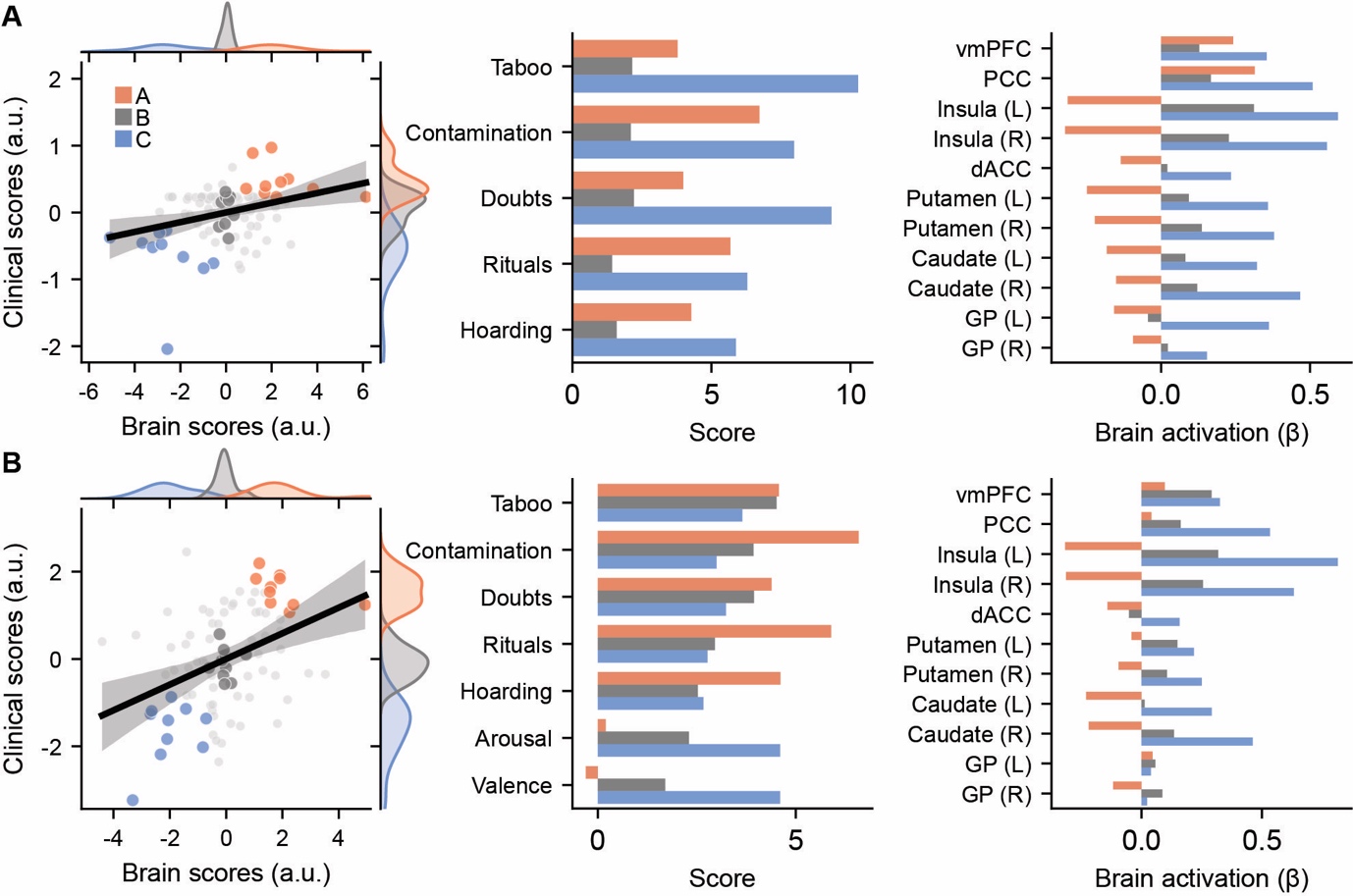

**Figure S7.** Partial least squares analysis with score-based groupings. **A.** Initial symptom factor model. The left-most panel demonstrates the relationship between individual brain activation PLSC scores and behavioural PLSC scores. The data has been divided into three groups with either high (orange, group A), neutral (grey, group B) or low (blue, group C) PLSC scores. Note these groups are not data driven but are for illustrative purposes only. The middle panel shows the average raw scores on the clinical data in reference to the three sub-groups. The right-most panel shows the average brain activations (beta coefficients) in reference to the three sub-groups. Note that these panels are complementary to Figure 4 in the manuscript which represent the correlation between patterns of PLSC scores and raw data. **B.** Secondary model with arousal and valence subjective ratings included. This plot is designed to aid in interpreting the findings and is not statistical in nature. The groupings shown are arbitrary.

### References

Abraham, A., Pedregosa, F., Eickenberg, M., Gervais, P., Mueller, A., Kossaifi, J., Gramfort, A., Thirion, B., & Varoquaux, G. (2014). Machine learning for neuroimaging with scikit-learn. *Frontiers in Neuroinformatics*, 14.

Avants, B. B., Epstein, C. L., Grossman, M., & Gee, J. C. (2008). Symmetric diffeomorphic image registration with cross-correlation: Evaluating automated labeling of elderly and neurodegenerative brain. *Medical Image Analysis*, *12*(1), 26–41.

Benjamini, Y., & Yekutieli, D. (2001). The Control of the False Discovery Rate in Multiple Testing under Dependency. *The Annals of Statistics*, *29*(4), 1165–1188.

Cox, R. W. (1996). AFNI: software for analysis and visualization of functional magnetic resonance neuroimages. *Computers and Biomedical Research*, *29*(3), 162–173.

Cox, R. W., & Hyde, J. S. (1997). Software tools for analysis and visualization of fMRI data. *NMR in Biomedicine: An International Journal Devoted to the Development and Application of Magnetic Resonance In Vivo*, *10*(4‐5), 171–178.

Dale, A. M., Fischl, B., & Sereno, M. I. (1999). Cortical surface-based analysis: I. Segmentation and surface reconstruction. *Neuroimage*, *9*(2), 179–194.

Eckart, C., & Young, G. (1936). The approximation of one matrix by another of lower rank. *Psychometrika*, *1*(3), 211–218.

Esteban, O., Blair, R., Markiewicz, C. J., Berleant, S. L., Moodie, C., Ma, F., Isik, A. I., Erramuzpe, A., Kent, M., & James, D. (2018). Fmriprep. *Software*.

Esteban, O., Markiewicz, C. J., Blair, R. W., Moodie, C. A., Isik, A. I., Erramuzpe, A., Kent, J. D., Goncalves, M., DuPre, E., Snyder, M., Oya, H., Ghosh, S. S., Wright, J., Durnez, J., Poldrack, R. A., & Gorgolewski, K. J. (2018). fMRIPrep: A robust preprocessing pipeline for functional MRI. *Nature Methods*, 1. https://doi.org/10.1038/s41592-018-0235-4

Foa, E. B., Huppert, J. D., Leiberg, S., Langner, R., Kichic, R., Hajcak, G., & Salkovskis, P. M. (2002). The Obsessive-Compulsive Inventory: Development and validation of a short version. *Psychological Assessment*, *14*(4), 485.

Fonov, V. S., Evans, A. C., McKinstry, R. C., Almli, C. R., & Collins, D. (2009). Unbiased nonlinear average age-appropriate brain templates from birth to adulthood. *NeuroImage*, *47*, S102.

Goodman, W. K., Price, L. H., Rasmussen, S. A., Mazure, C., Fleischmann, R. L., Hill, C. L., Heninger, G. R., & Charney, D. S. (1989). The Yale-Brown obsessive compulsive scale: I. Development, use, and reliability. *Archives of General Psychiatry*, *46*(11), 1006–1011.

Gorgolewski, K., Burns, C., Madison, C., Clark, D., Halchenko, Y., Waskom, M., & Ghosh, S. (2011). Nipype: A Flexible, Lightweight and Extensible Neuroimaging Data Processing Framework in Python. *Frontiers in Neuroinformatics*, *5*. https://www.frontiersin.org/articles/10.3389/fninf.2011.00013

Gorgolewski, K. J., Esteban, O., Markiewicz, C. J., Ziegler, E., Ellis, D. G., Notter, M. P., Jarecka, D., Johnson, H., Burns, C., & Manhães-Savio, A. (2018). Nipype. *Software*.

Greve, D. N., & Fischl, B. (2009). Accurate and robust brain image alignment using boundary-based registration. *Neuroimage*, *48*(1), 63–72.

Griffanti, L., Salimi-Khorshidi, G., Beckmann, C. F., Auerbach, E. J., Douaud, G., Sexton, C. E., Zsoldos, E., Ebmeier, K. P., Filippini, N., Mackay, C. E., Moeller, S., Xu, J., Yacoub, E., Baselli, G., Ugurbil, K., Miller, K. L., & Smith, S. M. (2014). ICA-based artefact removal and accelerated fMRI acquisition for improved resting state network imaging. *NeuroImage*, *95*, 232–247. https://doi.org/10.1016/j.neuroimage.2014.03.034

Hall, C. V., Harrison, B. J., Iyer, K. K., Savage, H. S., Zakrzewski, M., Simms, L. A., Radford-Smith, G., Moran, R. J., & Cocchi, L. (2022). Microbiota links to neural dynamics supporting threat processing. *Human Brain Mapping*, *43*(2), 733–749. https://doi.org/10.1002/hbm.25682

Hamilton, M. (1959). The assessment of anxiety states by rating. *British Journal of Medical Psychology*.

Jenkinson, M., Bannister, P., Brady, M., & Smith, S. (2002). Improved optimization for the robust and accurate linear registration and motion correction of brain images. *Neuroimage*, *17*(2), 825–841.

Katerberg, H., Delucchi, K. L., Stewart, S. E., Lochner, C., Denys, D. A. J. P., Stack, D. E., Andresen, J. M., Grant, J. E., Kim, S. W., Williams, K. A., den Boer, J. A., van Balkom, A. J. L. M., Smit, J. H., van Oppen, P., Polman, A., Jenike, M. A., Stein, D. J., Mathews, C. A., & Cath, D. C. (2010). Symptom Dimensions in OCD: Item-Level Factor Analysis and Heritability Estimates. *Behavior Genetics*, *40*(4), 505–517. https://doi.org/10.1007/s10519-010-9339-z

Klein, A., Ghosh, S. S., Bao, F. S., Giard, J., Häme, Y., Stavsky, E., Lee, N., Rossa, B., Reuter, M., & Chaibub Neto, E. (2017). Mindboggling morphometry of human brains. *PLoS Computational Biology*, *13*(2), e1005350.

McIntosh, A. R., & Lobaugh, N. J. (2004). Partial least squares analysis of neuroimaging data: Applications and advances. *NeuroImage*, *23*, S250–S263. https://doi.org/10.1016/j.neuroimage.2004.07.020

Mišić, B., Betzel, R. F., de Reus, M. A., van den Heuvel, M. P., Berman, M. G., McIntosh, A. R., & Sporns, O. (2016). Network-Level Structure-Function Relationships in Human Neocortex. *Cerebral Cortex*, *26*(7), 3285–3296. https://doi.org/10.1093/cercor/bhw089

Montgomery, S. A., & Åsberg, M. (1979). A new depression scale designed to be sensitive to change. *The British Journal of Psychiatry*, *134*(4), 382–389.

Obsessive Compulsive Cognitions Working Group. (2005). Psychometric validation of the obsessive belief questionnaire and interpretation of intrusions inventory—Part 2: Factor analyses and testing of a brief version. *Behaviour Research and Therapy*, *43*(11), 1527–1542.

Salimi-Khorshidi, G., Douaud, G., Beckmann, C. F., Glasser, M. F., Griffanti, L., & Smith, S. M. (2014). Automatic denoising of functional MRI data: Combining independent component analysis and hierarchical fusion of classifiers. *NeuroImage*, *90*, 449–468. https://doi.org/10.1016/j.neuroimage.2013.11.046

Tustison, N. J., Avants, B. B., Cook, P. A., Zheng, Y., Egan, A., Yushkevich, P. A., & Gee, J. C. (2010). N4ITK: improved N3 bias correction. *IEEE Transactions on Medical Imaging*, *29*(6), 1310–1320.

Zhang, Y., Brady, M., & Smith, S. (2001). Segmentation of brain MR images through a hidden Markov random field model and the expectation-maximization algorithm. *IEEE Transactions on Medical Imaging*, *20*(1), 45–57.

Zigmond, A. S., & Snaith, R. P. (1983). The hospital anxiety and depression scale. *Acta Psychiatrica Scandinavica*, *67*(6), 361–370.
